# Determinants of passive antibody efficacy in SARS-CoV-2 infection

**DOI:** 10.1101/2022.03.21.22272672

**Authors:** Eva Stadler, Khai Li Chai, Timothy E Schlub, Deborah Cromer, Mark N Polizzotto, Stephen J Kent, Claire Beecher, Heath White, Tari Turner, Nicole Skoetz, Lise Estcourt, Zoe K McQuilten, Erica M Wood, David S Khoury, Miles P Davenport

## Abstract

**Background:** A large number of studies have been carried out involving passive antibody administration for the treatment and prophylaxis of COVID-19 and have shown variable efficacy. However, the determinants of treatment effectiveness have not been identified. Here we aimed to aggregate all available data on randomised controlled trials of passive antibody treatment for COVID-19 to understand how the dose and timing affect treatment outcome.

**Methods:** We analysed published studies of passive antibody treatment from inception to 7 January 2022 that were identified after searching various databases such as MEDLINE, Pubmed, ClinicalTrials.gov. We extracted data on treatment, dose, disease stage at treatment, and effectiveness for different clinical outcomes from these studies. To compare administered antibody levels between different treatments, we used data on *in vitro* neutralisation of pseudovirus to normalise the administered dose of antibody. We used a mixed-effects regression model to understand the relationship between disease stage at treatment and effectiveness. We used a logistic model to analyse the relationship between administered antibody dose (normalised to the mean convalescent titre) and outcome, and to predict efficacy of antibodies against different Omicron subvariants.

**Findings:** We found that clinical stage at treatment was highly predictive of the effectiveness of both monoclonal antibodies and convalescent plasma therapy in preventing progression to subsequent stages (p<0.0001 and p=0.0089, respectively, chi-squared test). We also analysed the dose-response curve for passive antibody treatment of ambulant COVID-19 patients to prevent hospitalisation. Using this quantitative dose-response relationship, we predict that a number of existing monoclonal antibody treatment regimens should maintain clinical effectiveness in infection with currently circulating Omicron variants.

**Interpretation:** Early administration of passive antibody therapy is crucial to achieving high efficacy in preventing clinical progression. A dose-response curve was derived for passive antibody therapy administered to ambulant symptomatic subjects to prevent hospitalisation. For many of the monoclonal antibody regimens analysed, the administered doses are estimated to be between 7 and >1000 fold higher than necessary to achieve 90% of the maximal efficacy against the ancestral (Wuhan-like) virus. This suggests that a number of current treatments should maintain high efficacy against Omicron subvariants despite reduction in *in vitro* neutralisation potency. This work provides a framework for the rational assessment of future passive antibody prophylaxis and treatment strategies for COVID-19.

**Funding:** This work is supported by an Australian government Medical Research Future Fund awards GNT2002073 and MRF2005544 (to MPD, SJK), MRF2005760 (to MPD), an NHMRC program grant GNT1149990 (SJK and MPD), and the Victorian Government (SJK). SJK is supported by a NHMRC fellowship. DC, MPD, ZKM and EMW are supported by NHMRC Investigator grants and ZKM and EMW by an NHMRC Synergy grant (1189490). DSK is supported by a University of New South Wales fellowship. KLC is supported by PhD scholarships from Monash University, the Haematology Society of Australia and New Zealand and the Leukaemia Foundation. TT, HW and CB are members of the National COVID-19 Clinical Evidence Taskforce which is funded by the Australian Government Department of Health.

**Research in context:** *Evidence before this study:* We identified randomised controlled trials (RCTs) evaluating the effectiveness of SARS-CoV-2-specific neutralising monoclonal antibodies, hyperimmune immunoglobulin and convalescent plasma in the treatment of participants with a confirmed diagnosis of COVID-19 and in uninfected participants with or without potential exposure to SARS-CoV-2. The RCTs were identified from published searches conducted by the Cochrane Haematology living systematic review teams. A total of 37 randomised controlled trials (RCT) of passive antibody administration for COVID-19 were identified. This included 12 trials on monoclonal antibodies, 21 trials of convalescent plasma treatment, and 4 trials of hyperimmune globulin. These trials involved treatment of individuals either prophylactically or at different stages of infection including post-exposure prophylaxis, symptomatic infection, and hospitalisation. The level of antibody administered ranged from a 250 ml volume of convalescent plasma through to 8 grams of monoclonal antibodies. Data for analysis was extracted from the original publications including dose and antibody levels of antibody administered, disease stage and timing of administration, primary outcome of study and whether they reported on our prespecified outcomes of interest, which include protection against symptomatic infection, hospitalisation, need for invasive mechanical ventilation (IMV) and death (all-cause mortality at 30 days).

*Added value of this study:* Our study included data across all 37 RCTs of passive antibody interventions for COVID-19 and aggregated the studies by the stage of infection at initiation of treatment. We found that prophylactic administration or treatment in earlier stages of infection had significantly higher effectiveness than later treatment. We also estimated the dose-response relationship between administered antibody dose and protection from progression from symptomatic ambulant COVID-19 to hospitalisation. We used this relationship to predict the efficacy of different monoclonal antibody treatment regimes against the Omicron subvariants BA.1, BA.2, and BA.4/5. We also used this dose-response relationship to estimate the maximal efficacy of monoclonal antibody therapy in the context of pre-existing endogenous neutralising antibodies.

*Implications of all the available evidence:* This work identifies that both prophylactic therapy and treatment in the early stages of symptomatic infection can achieve significant protection from infection or hospitalisation respectively. The dose-response relationship provides a quantitative means to predict the change in efficacy of different monoclonal antibodies against new variants and in semi-immune populations based on *in vitro* neutralisation data. We predict a number of existing monoclonal antibodies will be effective for preventing severe outcomes when administered early in BA.4/5 infections. It is likely that these therapies will provide little protection in individuals with high levels of endogenous neutralising antibodies, such as healthy individuals who have recently received a third dose of an mRNA vaccine.

## Introduction

A large number of studies have investigated the effectiveness of passively administered antibodies, including convalescent plasma (CP), hyperimmune globulin (hIVIG) and monoclonal antibody (mAb) products. In particular, based on the high clinical efficacy reported for some of these products in randomised placebo-controlled trials (RCTs), multiple monoclonal therapies have received regulatory approval and recommendations for use in prophylaxis and / or treatment of COVID-19. However, these RCTs were carried out in primarily unvaccinated and SARS-CoV-2 infection-naïve individuals and occurred when ancestral-like SARS-CoV-2 variants were predominately circulating. The current major challenge for regulatory and clinical decision making is to predict the efficacy of individual monoclonal antibody products against new SARS-CoV-2 variants, as well as understanding whether and how these products should be used in a vaccinated or semi-immune population. At present these regulatory decisions have been based on *in vitro* neutralisation data, where loss of neutralising activity *in vitro* has been used to inform decisions for the withdrawal of emergency use authorisation for some antibodies against recent Omicron strains. However, to-date, there has been no data-informed method for determining how the loss of *in vitro* neutralising activity by a monoclonal is expected to impact the clinical efficacy of the product. Thus, a major challenge is to derive a quantitative method for predicting whether a given passive antibody regimen will remain effective against SARS-CoV-2 variants and/or in semi-immune populations.

In this study, we conducted a systematic search of the literature and aggregated data from 37 randomised controlled trials of passive antibody therapy to explore how the timing and *in vitro* neutralisation activity of different types of passive antibody treatments (including SARS-CoV-2-neutralising mAbs, convalescent plasma and hIVIG) predicts protection from COVID-19. To examine the impact of dose, we utilise a previously reported approach of normalising measures of antibody neutralisation from across multiple studies relative to the mean serum neutralisation titre of convalescent subjects^1^. This allows us to assess the effective dose of neutralising antibodies administered in different plasma and monoclonal antibody studies to determine the relationship between dose and efficacy. We demonstrate that across both CP and mAb studies there is a strong correlation between higher efficacy and treatment at earlier disease stage. Further we show a clear relationship between the dose of neutralising antibodies administered to ambulant COVID-19 patients and clinical efficacy in preventing hospitalisation. This quantitative relationship provides a clear mechanism for predicting the therapeutic efficacy of new and existing antibodies against variants of concern. We use this relationship to predict which antibodies are likely to maintain clinically relevant activity against the SARS-CoV-2 Omicron subvariants BA.1, BA.2, BA.4 and BA.5.

## Methods

### Search strategy and selection criteria for RCT of passive antibody therapy

We performed a systematic search of the literature to identify randomised controlled trials of passive antibody therapy for prevention and treatment of COVID-19. We included studies of SARS-CoV-2 neutralising mAbs, CP and hIVIG, in both individuals with a confirmed diagnosis of COVID-19 and individuals without confirmed diagnosis of COVID-19 (i.e. including pre-exposure, and peri-(post)-exposure stages). We excluded studies evaluating standard immunoglobulin and mAbs that were not specifically designed to target SARS-CoV-2 (**Fig. S1**). Details of the databases used to conduct the literature search, as well as details on the outcomes extracted from each study are reported in the supplementary methods.

### Data collection

Data for analysis was extracted from included studies, including dose and antibody levels of antibody administered, disease stage according to the latest WHO clinical progression scale, and timing of administration, primary outcome of study and whether they reported on our prespecified outcomes of interest, which include protection against symptomatic infection, hospitalisation, need for invasive mechanical ventilation (IMV) and death (all-cause mortality at 30 days) (summarised in **Table S1** and **Table S2**). We classified disease stage into the following categories – uninfected (pre-exposure, peri-(post-)exposure) or infected (symptomatic infection or hospitalised with moderate or severe disease). We also collected whether studies reported results for seronegativity/seropositivity of recipients at baseline.

### Data analysis and modelling

*Efficacy and 95% confidence interval*. We calculated the efficacy of preventing progression from one infection stage (including pre- and peri-(post-) exposure) to another stage using reported numbers of patients with disease progression and total numbers of patients in the following way:

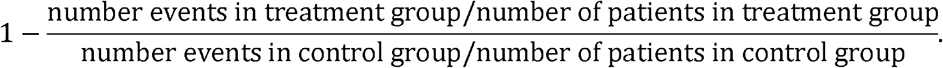

Thus, efficacy is 1 − “relative risk of progression”. We transformed the efficacy to percent and computed the corresponding 95% Confidence Interval (CI) as described in the supplementary methods.

#### Statistical analysis

Aggregated RCT data was analysed using a generalised linear mixed-effects modelling approach, with binomial error family and logarithmic link function (using R, version 3.6.0^2^, and the *glmer* function from the *lme4* package^3^). The model includes random intercepts for different trials to account for variability between different trials. We used this model to pool data from different studies and analyse the relationships between efficacy of mAb or CP/hIVIG treatment when used at different infection stages, to prevent different infection outcomes, and at different doses (**Fig. 1, Table S3, Table S4, Table S5, Table S6**, see supplementary methods for more details).

**Fig. 1.**
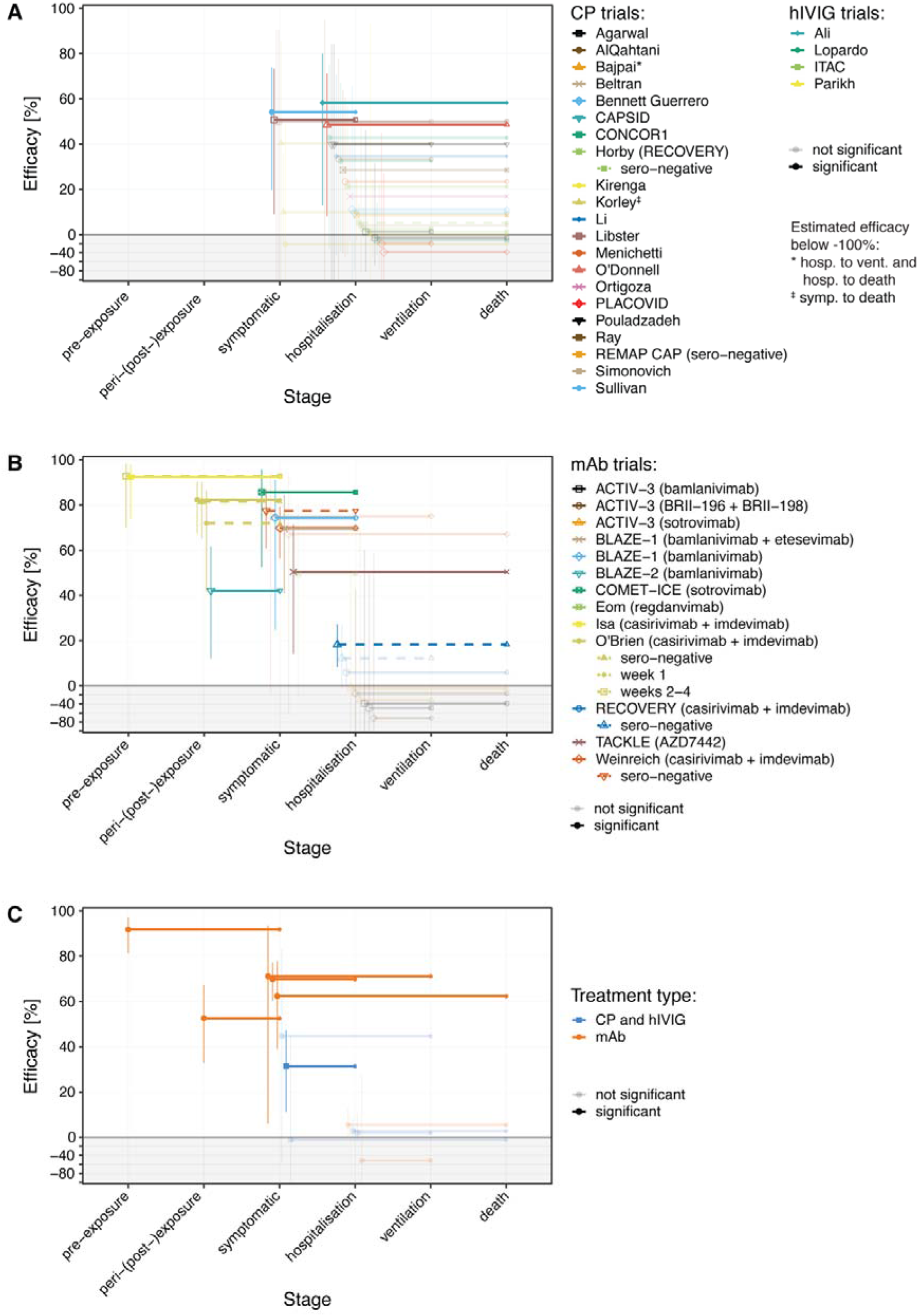
Effects of passive antibody treatment according to clinical stage at administration. The results for individual studies are indicated as lines, where the left-hand end of the line indicates stage at which antibodies were administered, and the right hand indicates the clinical outcome recorded. Some studies include multiple outcomes reported (shown as lines to different outcomes) or multiple subsets (shown as dashed lines). The y-axis shows efficacy for the indicated outcome (horizontal line position) and 95% confidence intervals for the efficacy (whiskers). Results for studies reporting negative efficacies are shown in the shaded region (note that y-axis is compressed here). (A) Protection observed in studies of plasma or hIVIG administration. (B) Studies involving administration of monoclonal antibodies. (C) Mean protection for treatment with monoclonal antibodies or convalescent plasma / hIVIG at different stages of infection and for different outcome stages (estimated using a generalised linear mixed-effects model, see supplementary methods).

#### Dose-response curve fitting

We used a maximum likelihood approach to fit a logistic function to the relationship between the administered dose of neutralising antibodies on the convalescent equivalence scale (estimated as described in the supplementary methods) and the efficacy reported in each RCT (**Fig. 2** and **Fig. S2**). Details of fitting approach and parameter estimation are described in the supplementary methods. All 95% confidence intervals for the fitted model and predictions from the model were performed using parametric bootstrapping (supplementary methods). All analysis was performed using the statistical software R (version 3.6.0)^2^.

**Fig. 2.**
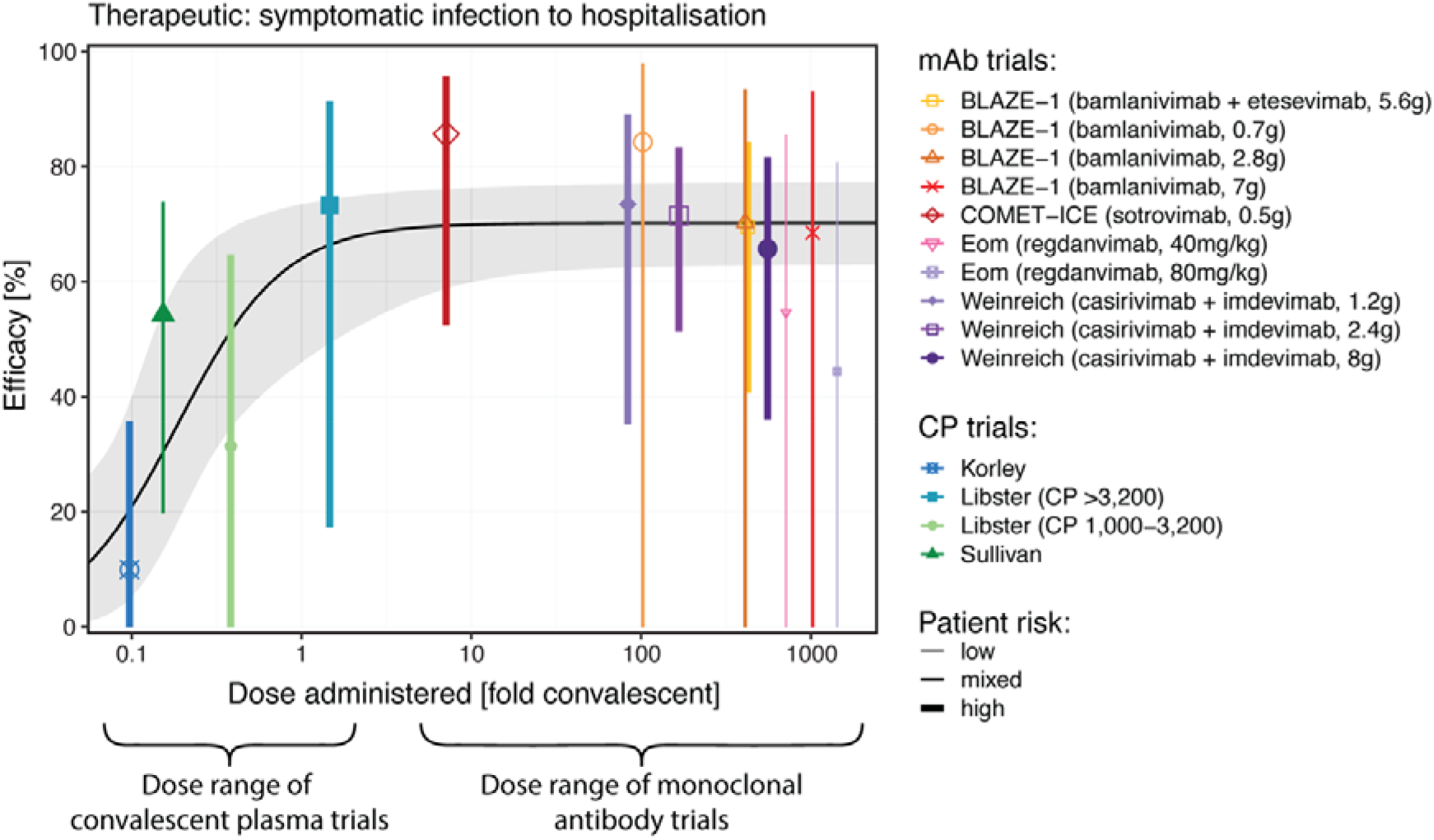
Estimating the dose-response curve for intravenous administration of neutralising antibodies protecting from progression from symptomatic infection to hospitalisation. The fitted curve allows for a maximum efficacy (estimated at 70.2%). The shaded area indicates the 95% confidence band.

#### Predicting efficacy against SARS-CoV-2 variants and in semi-immune individuals

The logistic model relating dose and efficacy was used to predict efficacy against variants, using a meta-analysis of the IC-50 of each monoclonal antibody against different SARS-CoV-2 variants. Similarly, for semi-immune individuals, we estimated the impact of pre-treatment circulating endogenous antibodies in reducing passive antibody treatment effect using the same logistic relationship between administered antibody dose and efficacy (supplementary methods). Confidence intervals of the predicted values were estimated by parametric bootstrapping (supplementary methods).

## Results

### Summary of included studies

We identified 37 RCTs evaluating passive antibody therapies for treatment or prevention of COVID-19 (**Fig. S1**). This included 21 evaluating convalescent plasma, four RCTs evaluating hIVIG and 12 primary studies of monoclonal antibodies (including eight different monoclonal antibodies or combinations). The studies varied in the protocol design (summarised in **Table S1** and **Table S2**), including stage of infection at the time of treatment, primary outcome measures, as well as the volume and antibody titres of the plasma administered. Specifically, all studies evaluating hIVIG products (four), 17 studies evaluating CP products and three studies evaluating monoclonal antibodies, were performed in hospitalised participants with moderate and/or severe disease and analysed efficacy in preventing invasive mechanical ventilation or death. Treatments were administered earlier and in milder infections (ambulatory patients, with mild disease) in three CP studies and five monoclonal antibody studies and were evaluated for efficacy to prevent progression to severe disease/hospitalisation. Two studies assessed monoclonal antibody efficacy as post-exposure prophylaxis administered to close contacts to prevent symptomatic infection (one of these studies also reported an efficacy in preventing infection beyond one week after administration^4^, which we assume was effectively true prophylaxis - since exposure most likely occurred after treatment). Finally, one study investigating true prophylaxis showed an efficacy of 92.4% in preventing symptomatic SARS-CoV-2 infection^5^. Thus, we grouped studies into broad categories by treatment (either CP/hIVIG or mAb), disease stage at enrolment, and outcomes reported and estimated the pooled efficacy by treatment group and outcome (**Fig. 1** and **Table S3**).

### Effects of timing on antibody efficacy

Despite the significant heterogeneity in trial design, clear patterns of decreasing efficacy with disease stage at treatment emerge (see **Fig. 1C** for the efficacy of mAb treatment (orange) and CP or hIVIG treatment (blue) by treatment and outcome stage). We tested this relationship using a generalised linear mixed-effects regression model and found that treatment at later disease stages (as an ordinal variable) was significantly associated with decreasing efficacy at preventing progression to the next stage (**Table S4** and **Table S5**), for both CP/hIVIG studies (for treated patients, the relative risk of progression increases by 1.42-fold per disease stage, 95% CI: 1.09–1.86, p=0.0089) and mAbs (relative risk 1.96, 95% CI: 1.70–2.28, p<0.0001). However, we note that this analysis aggregated studies with different disease outcomes, so we further tested this relationship with data stratified by outcome (i.e. for a given clinical outcome, testing whether earlier treatment was more effective). We found higher efficacy against a given clinical outcome was associated with earlier treatment for mAb studies (p<0.02 for all outcomes, **Table S4**) but no significant effect was observed in the CP/hIVIG studies (p=0.29 and 0.45 for outcomes IMV and death, respectively, **Table S5**). Thus, treatment either prophylactically or early in the course of infection is a major determinant to achieving protection with passive antibody administration in COVID-19.

### Dose-response for passive antibody administration

We next considered whether differences in efficacy between trials could be accounted for by the different doses and potencies of the products administered. Since there was generally no efficacy for passive antibody treatment in hospitalised individuals (**Fig. 1** and **Table S3**) and given the few studies of prophylaxis (**Fig. 1**), we focused only on treatments to prevent hospitalisation when treatment was administered to ambulant subjects with mild/moderate symptomatic infection (n=10 studies, two studies were excluded as they did not report hospitalisation as an outcome). Analysis of studies of prophylaxis and treatment in hospitalised subjects are described in the supplementary results. However, although one study analysed *in vivo* neutralisation capacity after treatment^6^, which provides a good surrogate measure of protection^1^, this was not routinely performed across the studies. Thus, it was necessary to estimate the effective dose of neutralising antibodies administered between the different studies. We have previously compared vaccine-induced neutralising antibody titres by reference to the ‘(geometric) mean convalescent titre’ seen in the first months after infection with the ancestral virus^1^. Therefore, we investigated whether a similar metric might be used to understand neutralising titres after passive antibody administration (supplementary methods). On this ‘convalescent equivalence’ scale, a group of individuals with a mean neutralising titre of 1-fold of convalescence has (on average) the same level of neutralising antibodies as the average convalescent individual (against ancestral virus after infection with ancestral virus). We find that most mAb studies administered doses of antibodies that would be equivalent to >100-fold the average neutralising titre observed in convalescent individuals (**Fig. 2** and **Table S7**). In **Fig. 2** we plot the ‘convalescent equivalent’ of different administered doses of mAbs and convalescent plasma against efficacy.

To derive a quantitative relationship between administered dose and protection from hospitalisation, we fitted this data using a logistic model allowing for a maximal efficacy (see methods and supplementary methods). We estimate a maximum protection of 70.2% (regardless of dose) and the administered dose to achieve 50% of this maximal effect (EC-50 for hospitalisation, equivalent to 35.1% protection overall) is 0.185-fold (95% CI: 0.087 – 0.395) the mean convalescent serum titre (**Table S8** and **Fig. S2**). We ensured this relationship was not overly affected by the results of any single study, by performing a sensitivity analysis using a ‘leave-one-out’ approach (**Fig. S3**). Of note, we observed a strong trend towards decreasing efficacy with higher administered doses, but this is not significant (p=0.054, mixed-effects logistic regression described in supplementary methods, **Table S9**). This dose-response relationship between administered dose and efficacy provides a direct quantitative means of predicting therapeutic efficacy based on *in vitro* neutralisation data.

### Predicting clinical protection from *in vitro* neutralisation of new therapies and variants

The analysis above compares antibodies by reference to their neutralisation titre against ancestral (Wuhan-like) virus. However, a number of Omicron sub-variants including Omicron BA.1, BA.2, BA.4 and BA.5 have been shown to have greatly reduced *in vitro* neutralisation by therapeutic monoclonal antibodies^7^. Thus, a major clinical question is which monoclonal antibody regimens are likely still effective against existing or emerging variants. The relationship between the administered neutralising dose and efficacy (**Fig. 2**) provides a quantitative means to predict the therapeutic efficacy of monoclonal antibodies against variants and under different dosing regimes, based solely on *in vitro* neutralisation data.

A systematic review of the literature on *in vitro* neutralisation of monoclonal antibodies against BA.1 and BA.2 was published previously^7^, in which the combined data on IC50s from all studies was made available. In addition, we conducted our own review of the literature to find all *in vitro* neutralisation data of monoclonal antibodies against BA.4/5 available up to 14 July 2022 (n=4 studies identified^8-11^, search strategy described in supplementary methods). Using these data we estimated the *in vitro* IC-50 of each antibody against the Omicron subvariants (**Fig. S4**). We found that several mAbs were predicted to maintain detectable *in vitro* neutralisation against individual Omicron subvariants, including bebtelovimab, cilgavimab/ tixagevimab and sotrovimab. Thus, we predicted (with confidence intervals) the therapeutic efficacy of these antibodies when administered to ambulant individuals in preventing progression to hospitalisation (supplementary methods, **Fig. 3**). The confidence intervals included uncertainty in the IC-50 estimates of antibodies against each variant (**Fig. S4** and **Fig. S5**) as well as uncertainty in the model parameters (**Fig. 2**). We find that the 600 mg cilgavimab/tixagevimab regimen (perhaps because of the tixagevimab component against BA.1 and the cilgavimab component against BA.2 and BA.4/5) is predicted to maintain >67.4% efficacy against the major Omicron subvariants (lower bound for the 95% CI for BA.4/5 is 34.8%) (**Fig. 3**). Also, 500 mg sotrovimab is predicted to have 69.0% (95% CI: 50.9-75.5), 63.0% (95% CI: 17.3-73.9) and 58.7% (95% CI: 6.17-73.4) efficacy against BA.1, BA.2, and BA.4/5, respectively. Since achieving a minimal efficacy is likely important to treatment decisions, we also estimated the confidence that a given treatment could achieve at least 30% efficacy in preventing hospitalisation. We estimate that tixagevimab/cilgavimab (600 mg) will have significantly higher than 30% protection against hospitalisation with BA.4/5 infection (p=0.018, **Fig. 3**). For sotrovimab, the predicted efficacy of a 500 mg dose was not significantly higher than 30% efficacy (p=0.15). However, the model predicts significant confidence that a 4-fold higher dose (2000 mg) of sotrovimab would provide significantly greater than 30% efficacy (p=0.024, **Fig. 3**). **Table S10** summarises the estimated efficacies against the Omicron variants.

**Fig. 3.**
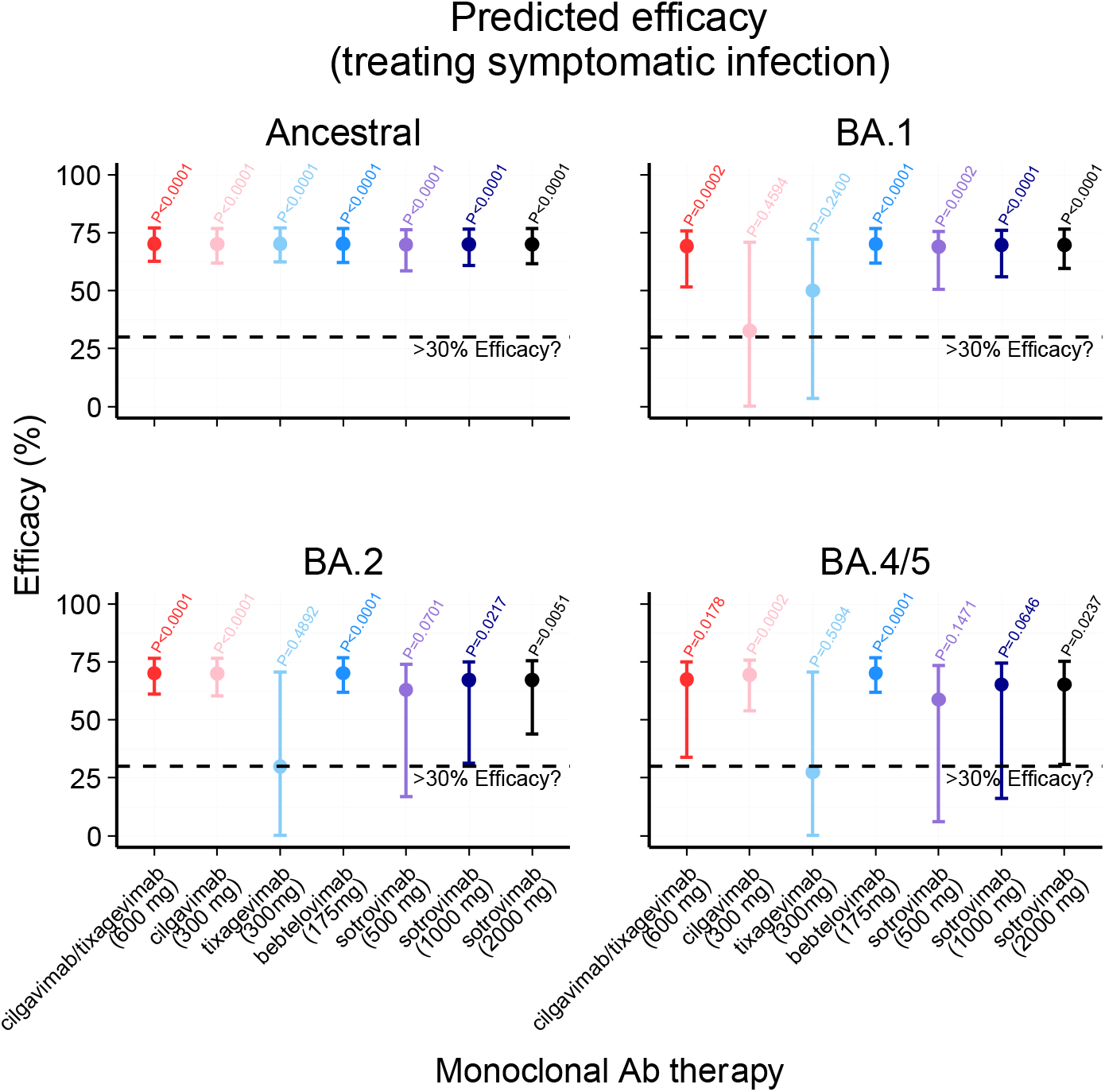
Predicted therapeutic efficacy of mAbs administered intravenously to ambulant COVID-19-positive subjects in preventing progression to hospitalisation. Predicted efficacy of cilgavimab/tixagevimab, cilgavimab, tixagevimab, bebtelovimab and sotrovimab at different doses against ancestral SARS-CoV-2 virus and the omicron variants BA.1, BA.2 and BA.4/5 is shown. The p-value indicates the confidence that the predicted therapeutic efficacy is at least 30% (for the efficacies and 95% CIs, see **Table S10**).

### Predicting clinical protection for semi-immune populations

The analysis and predicted efficacies above all consider passive antibody protection in immunologically naïve individuals. However, a major question is whether these therapies will still be effective in semi-immune (previously infected or vaccinated) individuals who have an endogenous neutralising antibody response. For example, if we assume that endogenous neutralising antibodies are similarly effective as administered monoclonal antibodies, then endogenous antibodies should already confer some degree of protection. If the endogenous neutralising antibody level already exceeds the predicted EC-90 for protection, for example, then adding more (monoclonal) antibodies may not improve protection. Thus, we used the relationship between dose and efficacy to predict the maximum efficacy of monoclonal antibody treatment when used in individuals with different levels of pre-existing endogenous neutralising antibodies (**Fig. 4**). Importantly, this analysis assumes that (a) a highly effective monoclonal is available against a particular variant, (b) endogenous antibodies (present before infection) provide at least the same level of protection as monoclonal antibodies administered in early infection, and (c) that neutralising antibodies are the only mechanism of vaccine protection (supplementary methods). For these reasons we might consider the resulting values as indicating a ‘theoretical maximum’ passive antibody protection for a given level of (pre-treatment) endogenous immunity.

**Fig. 4.**
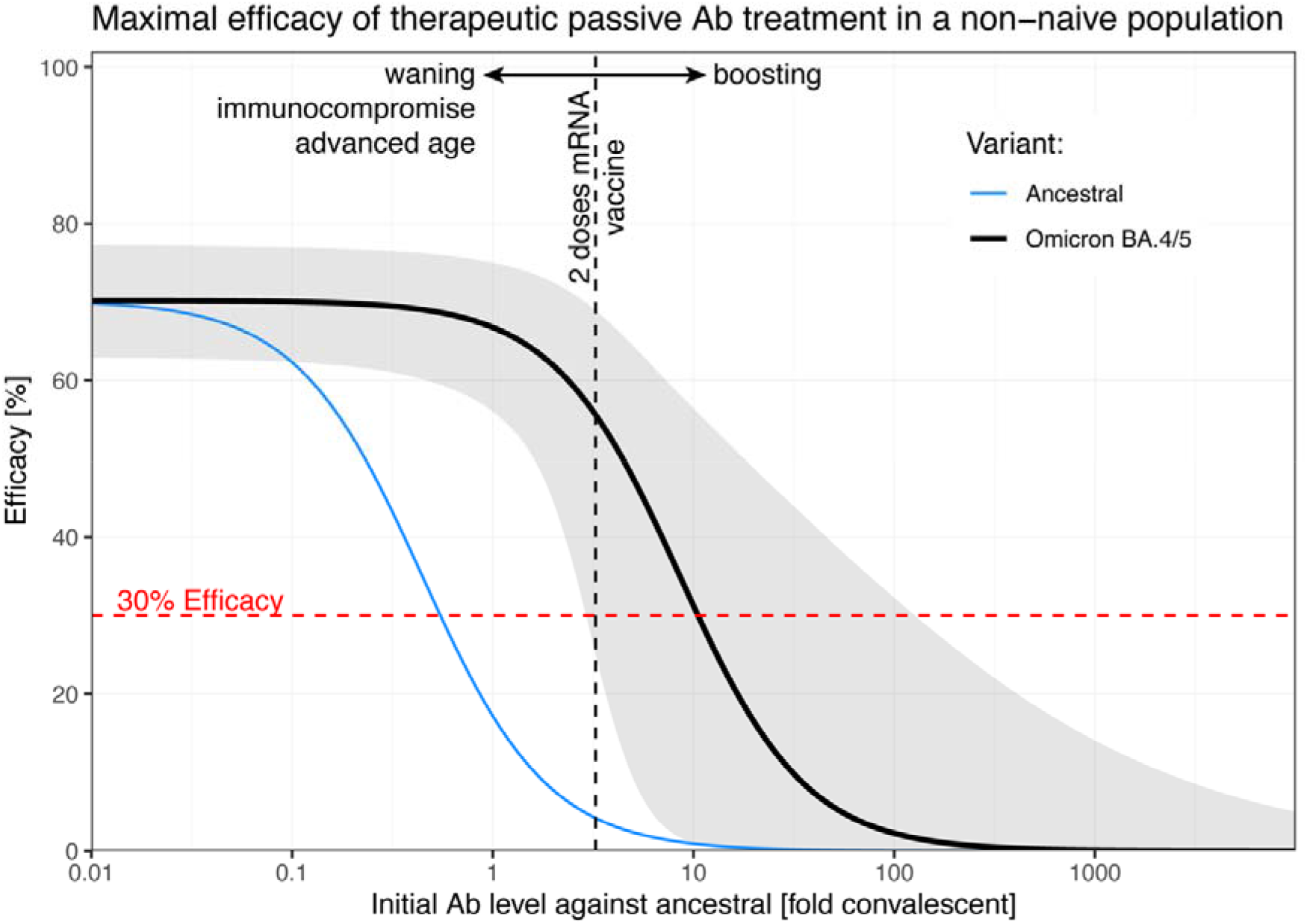
Predicted maximal efficacy of passive antibody treatment of symptomatic patients in a non-naïve population (assuming a fully effective antibody is available). Using the dose-response curve of therapeutic Ab treatment (**Fig. 2**), we predicted the theoretical maximal additional effect of passive Ab treatment of non-naïve patients (see supplementary methods) against ancestral (blue) and omicron variants BA.4/5 (black, with 95% confidence bands in grey). The mean neutralisation level shortly after two doses of an mRNA vaccine is indicated as a vertical black dashed line. This neutralisation level will decrease over time due to waning and is expected to be lower in advanced age or immunocompromise. Boosting is expected to increase endogenous neutralising antibody titres, leading to reduced maximal efficacy of passive antibody therapy.

As expected, at low endogenous antibody levels the maximum estimated benefit of monoclonal antibodies can be achieved (70.2% efficacy). However, after mRNA vaccination the endogenous neutralising antibody titres against the ancestral variant are sufficiently high that antibody treatment would not be expected to increase protection against the ancestral virus. However, vaccine-induced neutralising antibody titres are greatly reduced against the circulating Omicron variants (for example, Wang et al.^8^ report a 19.2-fold drop in neutralising antibody titres to the BA.4/5 variant in vaccinees). Combining this analysis of endogenous antibody titres to variants and the dose-response curve of passive antibody therapy, we predict the maximal potential efficacy of monoclonal antibodies in a recently mRNA vaccinated (2 doses) subject is reduced to 55.6% (95% CI: 25.0-68.9) (compared to 70.2% in naïve subjects). However, this maximal efficacy of neutralising antibody treatment is expected to be increased with waning immunity, advanced age, or immunosuppression (**Fig. 4**). Healthy immune individuals under 65, who have recently received a booster dose, are expected to have considerably higher endogenous neutralising responses^12^ and mAb efficacy is expected to be limited in these populations.

## Discussion

Despite the availability of effective vaccines for COVID-19, a significant population of elderly or immunosuppressed individuals may be unable to fully benefit from existing vaccines^13^. Passive antibody therapy has the potential to be used either prophylactically or therapeutically in this population. As we show in this meta-analysis, the available data from RCTs of passive antibody therapy demonstrates that both prophylactic therapy and treatment in the early stages of symptomatic infection can achieve significant protection from infection or hospitalisation, respectively. However, the continual emergence of variants and the loss of *in vitro* neutralisation activity of some mAbs has raised ongoing challenges for regulators and policy makers in deciding which monoclonal antibody treatments may gain or lose effectiveness against new circulating variants. These decisions have become reliant on *in vitro* measurements of monoclonal antibody neutralisation to predict *in vivo* efficacy. In some cases, the decision to maintain or discontinue treatment to a given variant may be relatively clear (if neutralisation is either undetectable or if the *in vitro* IC-50 remains unchanged). In the case of a partial loss of *in vitro* neutralisation effectiveness (a smaller change in IC-50), the decision may be more difficult. For example, sotrovimab was reported to show a 15.7-fold change in the *in vitro* EC-50 to the BA.2 variant, leading to the FDA withdrawing Emergency Use Authorisation for sotrovimab against BA.2 on 5 April 2022^14,15^. This highlights the importance of an evidence-based mechanism to predict how changes in *in vitro* potency will translate to clinical efficacy.

The relationship between the administered dose of a passive antibody therapy and clinical efficacy (**Fig. 2**) provides the first quantitative means to relate changes in *in vitro* neutralisation potency to outcomes from RCTs. Using this approach, changes in *in vitro* IC-50 can be used to directly predict *in vivo* efficacy. Unfortunately, even this calculation is not straightforward, as different studies often report different changes in IC-50 for the same antibody-variant combination. Tao et al.^7^ have comprehensively summarised the literature on neutralising potency of mAbs against the Omicron subvariants BA.1 and BA.2, and we have used this data to estimate the mean change in IC-50 and uncertainty in this estimate for different mAb-variant combinations. In addition to this, we added neutralisation data on BA.4 and BA.5 from four studies (**Fig. S4**). Using these *in vitro* data on the potency of each mAb against each variant, we predicted clinical efficacy of each mAb and found that at least some existing mAbs were expected to deliver greater than 30% efficacy against the Omicron subvariants (**Fig. 3**). A simple explanation for this is that many antibodies are currently administered at doses 10-1000-fold the predicted EC-90 against ancestral virus (**Fig. 2**). Thus, in some cases, a considerable increase in *in vitro* IC-50 can be tolerated before a given mAb loses clinical efficacy against a variant.

A major question is whether monoclonal antibody therapy will be useful in previously vaccinated or infected individuals who have already mounted an endogenous antibody response. Our analysis provides a clear prediction of a reduced efficacy of monoclonal antibody therapy with increasing endogenous antibody levels. However, it also suggests that the levels of neutralising antibodies to BA.4/5 after mRNA vaccination are not so high as to preclude a potential additional benefit of monoclonal antibody therapy. Importantly, we only estimate the theoretical maximum monoclonal antibody efficacy, which assumes that endogenous neutralising antibody responses provide the same protection as monoclonal antibody treatment. If endogenous non-neutralising responses or cellular immune responses provide a higher level of protection than estimated, this will obviously reduce the benefit of monoclonal antibody treatment. Interestingly, one study has suggested that treatment of hospitalised subjects may still be effective in seronegative subjects but not in seropositive individuals^16^, supporting the idea of a trade-off between endogenous and therapeutic antibody effects.

We also found that the timing of antibody administration is a major factor in determining the success of treatment. For example, for the same antibody doses we see >90% protection from symptomatic infection when given as prophylaxis^5^, but only ≈70% protection from progression to hospitalisation when given to ambulant COVID-19 patients^17^, and negligible protection when given to hospitalised subjects^16^. A major question is whether treatment even earlier after symptom onset might produce better outcomes? For example, would treatment on day 1 versus day 5 post onset of symptoms have higher effectiveness? This was not clear from the studies summarised in this paper (**Fig. S6**), but in an RCT on the therapeutic efficacy of cilgavimab/tixagevimab (published after our meta-analysis) a clear decline in efficacy was observed with later treatment^18^. Interestingly, despite their very different mode of action, studies of antiviral treatment also suggest that early treatment (prior to hospitalisation) is important^19,20^. Studies are urgently needed to identify the optimal time for passive antibody treatment in symptomatic subjects.

This study has a number of limitations. Firstly, it aggregates studies using different therapeutics and with different enrolment and outcome criteria. Secondly, it tries to equate administered doses of convalescent plasma and monoclonal antibodies based on a single study comparing *in vitro* neutralisation of pseudovirus^21^ (although this study reports IC-50s that are relatively consistent with the meta-analysis reported by Tao et al.^7^, **Fig. S4**). In the case of convalescent plasma, we consider mean titre of donor plasma and mean plasma volume for dilution, which does not reflect the considerable variability that exists between individual donor plasma neutralisation titres and individual recipients’ plasma volumes. In addition, we consider only the ‘administered dose’, as the studies did not directly measure plasma neutralisation titres in recipients after administration. When predicting the efficacy of therapies against different variants and in semi-immune populations, we assume the relationship between efficacy and dose continues to hold in these scenarios. Importantly, the dose-response curve for passive antibody administration is parameterised from studies of intravenous administration, where peak antibody levels are achieved very rapidly (**Fig. 2**). However, studies show a significant delay in achieving peak serum antibody concentrations after intramuscular administration^22^. Given the importance of timing of administration on the effectiveness of mAbs (**Fig. 1**), a delay in achieving an effective circulating antibody concentration might be expected to lead to lower efficacy. This may be particularly relevant for the use of cilgavimab/tixagevimab, which is currently administered both intramuscularly and intravenously^18,23^. Clinical studies to confirm that the dose-response curve is predictive of protection against variants of concern (VOCs) and in semi-immune populations are urgently required. Despite these limitations, a major strength of our approach is to apply a rigorous quantitative analysis to relate in vitro neutralisation potency with clinical outcomes based on the available data on passive antibody treatment for SARS-CoV-2. Such an evidence-based framework is crucial to inform clinical practice at a time when it has become necessary to rely on *in vitro* data to predict therapeutic efficacy.

This work provides quantitative and testable predictions of how passive antibody therapy may be optimally deployed to benefit a larger number of subjects. Further work is clearly required to better understand the relationship between administered dose and *in vivo* neutralising antibody titres and the impact of different SARS-CoV-2 variants on antibody efficacy. This work provides a quantitative framework to help guide rational decisions for the deployment of this important class of therapeutics.

## Data Availability

All data and code will be made available on GitHub upon publication.

## Ethics statement

This work was approved under the UNSW Sydney Human Research Ethics Committee (approval HC200242).

## Data and code availability

All data and code will be made available on GitHub upon publication.

## Acknowledgements

The authors thank all participants and original study teams of the studies analysed. This work would not have been possible without the data from these studies. We also thank David Sullivan for his help in understanding antibody titres measured in Sullivan et al.^24^.

We thank the authors from the Cochrane living systematic review teams of ‘Convalescent plasma or hyperimmune immunoglobulin for people with COVID-19: a living systematic review’ and ‘SARS-CoV-2-neutralising monoclonal antibodies for treatment of COVID-19’ and Steve McDonald for their contribution in developing the search strategies and identification of the included trials. We also thank the National COVID-19 Clinical Evidence Taskforce members for helpful discussions.

## Author contributions

ES, KLC, DC, SJK, EMW, ZKM, DSK and MPD contributed to conceptualization, supervision, and resources. ES, KLC, DC, NS, LE, EMW, ZKM, CB, HW, TT, DSK and MPD contributed to data curation, methodology, formal analysis, and visualization. All authors contributed to the writing and reviewed and approved the final report. All authors had full access to all the data in the study and had final responsibility for the decision to submit for publication.

