## Supplementary Material for "Determinants of passive antibody efficacy in SARS-CoV-2 infection"

#### Supplementary methods

##### Outcomes from each study that were included from systematic literature search

We included the following outcomes in all participants:

- All-cause mortality at day 30 or last follow up
- Need for invasive mechanical ventilation

We included the additional outcomes in participants with asymptomatic or mild disease:

- Admission to hospital

We included the additional outcomes in participants at the pre-exposure, peri-exposure or post-exposure stage without confirmed diagnosis of COVID-19:

- Infection
- Symptomatic infection
- Admission to hospital

We explored subgroups, including low vs. high titre CP and serostatus of patients at baseline (seronegative vs. seropositive).

We identified studies from published search strategies conducted by the Cochrane Haematology living systematic review teams which searched the following databases - MEDLINE, Embase, the Cochrane COVID-19 Study Register, Pubmed, Epistemonikos L*OVE List Coronavirus disease, World Health Organization COVID-19 Global literature on coronavirus disease and trial registry platforms, including ClinicalTrials.gov, WHO International Clinical Trials Registry Platform (ICTRP) and medRxiv (see references[1,2]). Detailed information about the search strategies used are available in published versions of the Cochrane living systematic review appendices. Additional recent studies were identified from review articles, monitoring of ongoing trials and ongoing updated searches until 7 January 2022.

##### Analysis of efficacy by treatment stage

To visualize the efficacy by treatment stage and outcome stage for each trial, we used the following stages for all trials:

- pre-exposure,
- peri-(post-)exposure,
- symptomatic,
- hospitalisation,
- ventilation, and
- death.

If a trial contained treatment with more than one monoclonal antibody, we considered different antibody treatments separately (e.g. ACTIV-3 sotrovimab treatment group and ACTIV-3 BRII-196 + BRII-198 treatment group). However, if a trial contained multiple doses of the same antibody treatment, then we pooled the different doses by adding the number of events and individuals in the different treatment and control groups, respectively.

For the O'Brien, RECOVERY, and Weinreich trials, data for patients who were seronegative at baseline were available and these were included as a subgroup in **Fig. 1A**, **B** (dashed horizontal lines).

We calculated efficacy of preventing a stage transition calculated in the following way:

$$1 - \frac{\text{number events in treatment group/number of patients in treatment group}}{\text{number events in control group/number of patients in control group}}$$

with 95% Confidence Interval (CI):

$$1 - \exp\left(\log\left(\frac{e_t/n_t}{e_c/n_c}\right) \pm 1.96 \times \sqrt{\frac{n_t - e_t}{n_t \times e_t} + \frac{n_c - e_c}{n_c \times e_c}}\right),$$

where $e_t$ and $n_t$ denote the number of events and total number of patients in the treatment group and $e_c$ and $n_c$ denote the number of events and total number of patients in the control group, respectively. This formula for the 95% CI assumes that the natural logarithm of the risk ratio is approximately normally distributed. Note that if there are zero events in either the treatment or the control group, then efficacy would be 100% or would be undefined (and similarly the 95% CI could not be computed using the above formula). We decided to omit one study with zero cases in the treatment group[3], as zero events often either indicated a low sample size or high uncertainty. The results are visualized in **Fig. 1A** and **B**.

**Pooling data from different trials**

To look at the average effect for different treatments and stages (**Fig. 1C**), we pooled data from different trials to visualize the efficacy of monoclonal antibody or plasma treatment by stage. We classified trials using the same treatment and outcome stages as described above (pre-exposure, peri-(post-)exposure, symptomatic, hospitalisation, ventilation, and death) and pooled different trials with the same treatment type (mAb or plasma/hIVIG) and same stage transitions (e.g. symptomatic to hospitalisation). For the O'Brien trial, we included the data in the two subgroups of infection within one week of treatment (the treatment stage for this group is peri-(post-)exposure) and infections beyond one week, i.e. two to four weeks, after treatment (for this subgroup, we assumed the treatment stage to be pre-exposure). For all other trials no subgroups were used (except for ACTIV-3 that was always analysed separately for sotrovimab and BRII-196 + BRII-198 treatment groups).

To account for differences between trials such as differences in the monoclonal antibody administered, patients' risk for progression to severe disease, or the time and location the trials where conducted, we used a mixed-effects logistic regression model with random intercepts for different trials. We used custom algorithms in the R software package (version 3.6.0)[4]. To compute the average efficacy for different treatments at different stages we used the function *glmer* from the lme4 package[5] with the binomial distribution and link function "*log*". Profile likelihood 95% CIs were computed using the "*confint*" function. We then transformed the relative risk and the corresponding 95% CI to efficacy (= 1- relative risk). The results are visualized in **Fig. 1C**.

**The effect of the initial stage on efficacy**

To investigate whether treatment at an earlier stage is more efficacious than at a later stage, we considered treatment with monoclonal antibodies and with plasma-products (CP or hIVIG) separately to take into account the difference in administered doses between these treatment types (**Fig. 2**). The initial stage is the stage at which patients were treated. As for the pooling of data from different trials, we used a mixed-effects logistic regression model with random intercepts for different trials and also included the variables treatment, initial stage, and an interaction term between treatment and the initial stage. The R[4] function *glmer* from the lme4 package[5] with the binomial distribution and link function "*log*" was used to compute relative risks, profile likelihood 95% CIs were computed using the "*confint*" function, and the significance of variables was tested using a chi-squared test with the function "*drop1*".

We considered two different ways of analysing the effect of the treatment stage on efficacy.
First, we considered progression to the next stage. We considered only the following stage transitions:

- pre-exposure to symptomatic,
- peri-(post-)exposure to symptomatic,
- symptomatic to hospitalisation, and
- hospitalisation to death

We excluded the stage “ventilation” because more data was available on “hospitalisation” and “death” and there were no transitions from “ventilation” to “death” in the data. To quantify the treatment effect by initial stage, we converted the initial stage to a continuous numerical variable starting from pre-exposure (transformed to 0) through to hospitalisation (transformed to 3). For both mAb and plasma treatment, the interaction between treatment and treatment stage was significant with a higher relative risk (i.e. lower efficacy) for treatment at a later stage.
Second, we considered the effect of the treatment stage for each outcome separately, i.e. for a chosen outcome stage (e.g. symptomatic infection) we compared the treatment effect for different treatment stages (e.g. comparing pre-exposure treatment and peri-(post-)exposure treatment in preventing symptomatic infection). For each outcome stage, there were maximally two disease stages at treatment in the data, so we used treatment stage as a categorical variable for this analysis. Note that for the outcome “hospitalisation” the only treatment stage was “symptomatic” and we could thus not compare the effect of different treatment on hospitalisations. For mAb treatment, treating at an earlier stage was significantly better than at a later stage for all outcomes. For plasma treatment (CP and hIVIG), there was no significant interaction between treatment and treatment stage for the outcomes ventilation and death (note that there is also no significant effect of treatment for these outcomes for all convalescent plasma treatment studies combined, see **Fig. 1C**). The results can be found in **Tables S4** and **S5**.

**Estimating the administered neutralising antibody dose relative to titres in convalescent individuals**

Previous analyses have compared neutralisation titre after vaccination by normalising to the ‘mean early convalescent titre’ (after COVID-19)[6,7]. Thus, to compare the efficacy of passive antibody treatments with different doses of neutralising antibodies, we normalized the dose relative to the ‘mean convalescent titre’ in the first months after infection.
For convalescent plasma treatment, if unselected convalescent plasma is used, we might consider the product itself to have a mean titre of ‘1-fold’ of the average convalescent serum titre, and the average ‘titre administered’ to reflect simply the plasma dilution upon administration (e.g. if a volume of 250mL of unselected convalescent plasma were administered to a recipient with 3L plasma volume, we could estimate that the equivalent of 0.083-fold the mean convalescent titre was administered). Importantly, we considered the mean administered dose, not implying the neutralisation titre of recipient plasma (if measured directly) would necessarily reflect this, since this estimate ignores any potential loss of product or loss of potency after administration. Similarly, some convalescent plasma studies specified that high titre plasma were selected; where this was the case, we normalised using the available data (**Table S7**). For this analysis, we focused on trials that treated outpatients and reported hospitalisations as an outcome, i.e. the convalescent plasma studies by Libster et al[8], Korley et al[9], and Sullivan et al[10].
Comparison of the effective neutralisation titres of different monoclonal antibody treatments is more challenging since equating this to the mean convalescent titre is not straightforward. McCallum et al[11] investigated both convalescent serum neutralisation and the *in vitro* IC-50 of different mAb in

the same SARS-CoV-2 pseudotyped virus neutralisation assay (see Fig. 2L and Fig. 3C of the original study)[11]. We assumed a mean plasma volume of 3 litres or 45mL/kg to convert the antibody dose to a mean antibody concentration. Dividing the antibody concentration by the respective antibody's IC-50, we obtained the fold-IC-50 for each antibody and each dose. We then calculated the dose in fold-convalescent by noting that the average convalescent subject in this study had 347.6 times the ID-50 (as extracted from McCallum et al) in their plasma, and that each mAb was administered at various levels (fold of the IC-50, **Table S11**). Thus, the fold-convalescent level for each mAb at each dose was estimated by dividing the fold-IC-50 by the mean convalescent neutralization titre of 347.6 (**Table S11**). For antibody combination treatments, we used the maximum of each individual antibody's fold-convalescent dose. The resulting fold-convalescent doses for each monoclonal antibody trial and each dose used can be found in **Table S7**.

Importantly, for both convalescent plasma and monoclonal antibodies we assume 100% distribution of the antibody in plasma, ignoring any potential loss during injection or thereafter. If there is a loss of antibody during infusion or thereafter this would lead to a tendency for reported administered titres to be higher than the serum titre that might be directly measured *in vivo* after infusion. Gordon et al[12] studied the pharmacokinetics of convalescent plasma after administration of a volume of 5 ml/kg to infants (leading to an estimated dilution of approximately 10-fold[13]). Direct measurement of recipient titres 30 minutes after infusion found a mean of 6.2% of donor titres, suggesting a decrease in titre of around 40% compared to what would be predicted by dilution alone. Thus, there may be a tendency for the 'administered dose' to be higher than a serum neutralisation level that might be directly measured *in vivo* after treatment.

**Estimating the mAb dose in mg for different efficacies in preventing hospitalisation**

To calculate the mAb dose of monoclonal antibodies that is required for different efficacies of preventing hospitalisation, we used the same approach. For each antibody we computed the dose for the EC-50, and EC-90 using the following formula:

$$\text{dose [mg]} = \text{dose [fold conv.]} \times \text{neutr}_{\text{conv.}} \times \text{IC}_{50} \left[\frac{\text{mg}}{\text{L}}\right] \times \text{total plasma volume [L]} \qquad \text{(Eq. 1)}$$

where dose [fold convalescent] was obtained from the dose response curve for prevention of hospitalisation (corresponding to the EC-50 or EC-90 efficacy, respectively), the neutralization titre of the convalescent is 347.6 (as extracted from McCallum et al[11]), the IC-50's of each antibody were extracted from McCallum et al[11] and can be found in **Table S11**, and we assumed again a mean total plasma volume of 3 litres.

**Estimating the administered dose fold-convalescent for CP trials**

Similar to the computation of the fold-convalescent dose of monoclonal antibody studies, we also calculated the fold-convalescent dose of three trials that treated symptomatic patients with convalescent plasma and reported hospitalisation as an outcome. These trials were published by Korley et al[9], Libster et al[8], and Sullivan et al[10].

Korley et al treated patients with 250mL of convalescent plasma with a median ID50 of 641 as measured with the PRNT assay as described by the Broad Institute[9]. Di Germanio et al reported convalescent neutralization using the same assay[14]. We found the geometric mean of convalescent to be 551 (data extracted from Fig. 2B of Di Germanio et al[14], first time point for each individual). As before, we assume a plasma volume of 3 litres to calculate the fold-convalescent dose of 0.097 for the Korley et al trial.

To find the dose administered in terms of fold-convalescent for the trial by Libster et al[8], we extracted the distribution of IgG titres in plasma donors from Figure S4. If we assume that the 72 extracted titres are from 72 subjects that represent the top 28th centile[8], then these must have been drawn from a theoretical population of 257 donors. Thus, we estimated that 185 individuals excluded from plasma donations for the trial by Libster et al because their titre was below the threshold of 1,000. With the extracted distribution of titres above the threshold of 1,000 and the number of excluded donors with titres below 1,000, we estimated the distribution of titres by fitting a normal distribution to the $\log_{10}$-transformed titres. We used a maximum likelihood approach with the following loglikelihood function:

$$l(p_1, p_2) = \sum \log\big(N(\log_{10}(t_{extracted}), p_1, \exp(p_2))\big) + n_{excluded} \times \log\big(\Phi(\log_{10}(1{,}000), p_1, \exp(p_2))\big),$$

where $p_1$ and $p_2$ are the parameters of the normal distribution, $N$ and $\Phi$ denote the probability density function and the cumulative distribution function of the normal distribution, respectively, $t_{extracted}$ are the extracted titers, and $n_{excluded}$ is the number of individuals excluded due to a titre below the threshold of 1,000. We obtained a mean convalescent titre of 360 (95% CI: 239 to 542). The volume of convalescent plasma used by Libster et al was 250mL and we assumed again a total recipient plasma volume of 3 litres. As Libster et al provided primary end point outcomes for recipient of donor plasma above or below the median of 3,200, we also computed the fold convalescent for these two groups separately. For the first group, who received a donor plasma with a titre between 1,000 and 3,200, we estimated the median donor titre to be 1,653 using our fitted distribution. With a dilution of 250mL in the 3 litres of recipient plasma and the median convalescent titre 360, this group has an administered dose of approximately 0.38-fold convalescent. For the second group, treated with plasma with a titre above 3,200, the median titre was estimated at 6,330 giving a dose administered of 1.47-fold convalescent.

Sullivan et al selected plasma with the highest 60 to 70% titres from unselected donors (personal communication, February 22 and 25, 2022). We assume that the $\log_{10}$ titres follow a normal distribution (with mean 0 which corresponds to 1 dose in fold-convalescent and standard deviation 0.46[15]). Sampling 1,000,000 times from this distribution and selecting the top 65% of titres, we find that the geometric mean of the top 65% of titres is 1.84-fold convalescent. With a dilution of 250mL of donor plasma in 3 litres of total recipient plasma, the administered dose in the trial by Sullivan et al is 0.153-fold convalescent.

**Dose-response curve fitting**

We fitted a logistic efficacy function to the dose and efficacy data for prevention of hospitalisation after treatment of symptomatic patients to obtain a relationship between the administered dose and the level of protection.

The dose and efficacy data were extracted from mAb and CP trials with the dose converted to fold convalescent as described above. We used an efficacy function that is logistic function of $\log_{10}$-transformed doses, i.e.

$$E(d \mid m, g, d_{half}) = \frac{m}{1 + \exp\left(-g\left(\log_{10}(d) - \log_{10}(d_{half})\right)\right)},$$

where $d$ denotes the dose, $m$ the maximal efficacy, $g$ the steepness of the curve, and $d_{half}$ the dose at which the half-maximal efficacy is achieved. We fitted this efficacy function to the count data from individual studies (i.e.: we used the total number of subjects and number of events in each of the treatment and control groups, rather than simply fitting to the summary statistic of 'efficacy'). This allowed us to take into account the variance in the number of participants in different trials and thus the uncertainty in the efficacy estimates. We used a maximum likelihood approach with the following likelihood function:

$$L(p) = \prod_{trials} \mathrm{Binom}(h_c, n_c, b) \times \mathrm{Binom}\left(h_t, n_t, b \times \left(1 - E(d \mid m, g, d_{half})\right)\right),$$

where $p$ denotes the parameters of the likelihood function, i.e. the three parameters of the efficacy function $(m, g, d_{half})$ and the baseline risk $b$ for each trial, Binom is the probability mass function of the binomial distribution, and for each trial, $h_c$ and $n_c$ are the numbers of hospitalisations and total number of individuals in the control group, $h_t$ and $n_t$ are the numbers of hospitalisations and total number of individuals in the treatment group, and $b$ is the baseline risk which is reduced by the efficacy of treatment for the treatment group. The initial values of the trial baseline risk for optimization were $h_c/n_c$.
We used parametric bootstrapping to compute the 95% confidence region for the efficacy function by sampling the three parameters for the efficacy function 100,000 times using the *rmvnorm* function (with mean and covariance matrix from the parameter estimate) from the *mvtnorm* package[16,17] in R (version 3.6.0)[4]. For each dose, the 95% confidence region was then computed using quantiles of the efficacy functions with the different sampled parameter values. The resulting estimated efficacy function and 95% confidence region is shown in **Fig. 2** and **Fig. S2**. The 95% CIs for the EC-50 and EC-90 doses were computed similarly, by bootstrapping the slope parameter and the EC-50 dose 100,000 times but using the estimated maximal efficacy (instead of also bootstrapping the maximal efficacy parameter, **Table S8**).

To study the sensitivity of the results to individual trials, we performed a leave-one-out analysis. We fitted the dose response curve again as described above to the data omitting one study at a time. The resulting parameter values for the fit to all data as well as to the data omitting one study are visualized in **Fig. S3**. We find that the estimates of the maximal efficacy are consistent at about 70% (range: 68.7 to 72.1%). The half-maximal dose (EC-50 dose) estimates vary between 0.069- and 0.38-fold convalescent which agrees well with the 95% CI of the fit to all data (0.087 to 0.395). Omitting the Korley trial had the greatest impact on the EC- 50 (0.069). The slope parameter estimates also agree with the 95% CI of the fit to all data (1.09 to 9.33), except if the Libster trial is left out (15.29).

To investigate the efficacy in preventing death after mAb treatment of hospitalised patients, we also fitted a dose response curve to data for the mAb treatment of hospitalised patients. We used the same approach as described above. As the only significant efficacy in this subset of the data is for the treatment of sero-negative patients and patients with unknown sero-status in the RECOVERY trial, we considered the sero-status subgroups of the RECOVERY trial in this analysis. The resulting fit is shown in **Fig. S8**.

**Fold increase of hospitalisations by administered mAb dose**
We found that monoclonal antibodies were administered at 7.9- to more than 1000-fold the EC-90 dose (**Table S12**). Thus, we aimed to investigate how the number of hospitalisations that are averted

by mAb treatment changes if the dose is reduced and if the antibody is a limited product. For a population-at-risk of $N$ individuals with risk of hospitalisation $r$ and $n_t$ treated individuals that are treated with a dose $d$ and corresponding efficacy $eff(d)$, the number of hospitalisations averted by treatment is given by

$$N \times r - \left[(N - n_t) \times r \; + \; n_t \times r \times \left(1 - eff(d)\right)\right] = \; n_t \times r \times eff(d).$$

If the administered dose $d$ is reduced to a new dose $d_{new}$, then the number of patients that can be treated increases to $n_t \times (d/d_{new})$ but the efficacy reduces from $eff(d)$ to $eff(d_{new})$. Overall, the number of hospitalisations averted under the treatment strategy with the new, lower dose compared to the current treatment strategy is given by

$$\frac{n_t \times (d/d_{new}) \times r \times eff(d_{new})}{n_t \times r \times eff(d)} = \frac{d \times eff(d_{new})}{d_{new} \times eff(d)}.$$

We visualised this fold increase in the number of hospitalisations averted with a lower dose treatment using our fitted logistic efficacy function and the currently administered doses of various mAbs (**Fig. S9**). For mAbs that are administered at various doses, we show the fold increase of hospitalisations averted for the lowest dose currently used. We also computed the fold increase in hospitalisations averted for treatment with the EC-90 dose compared to the lowest dose currently used, the maximal fold increase in hospitalisations averted, and the dose (in milligram) for the maximal fold increase in hospitalisations averted (**Table S13**).

**Efficacy of preventing hospitalisation by administered mAb dose**

The efficacy of preventing hospitalisation appears to be decreasing with an increasing administered dose of monoclonal antibody, however there is also high uncertainty in the efficacy estimates as indicated by big confidence intervals (**Fig. 2**). To quantify the effect of administered mAb dose on the efficacy of preventing hospitalisations, we used a mixed-effects logistic regression model. The model contains random intercepts for different trials, a treatment variable ("treatment" or "control"), the $\log_{10}$-transformed administered dose, the patient risk for progression to severe disease ("low", "mixed", or "high"), and the interaction of the $\log_{10}$-transformed administered dose and treatment. The R[4] function *glmer* from the lme4 package[5] with the binomial distribution and link function "*log*" (a log-binomial regression model) was used to compute relative risks, profile 95% CIs were computed using the "*confint*" function, and the significance of the patient risk and the interaction of the $\log_{10}$-transformed administered dose and treatment was tested using a chi-squared test with the function "*drop1*". The results can be found in **Table S9** and indicate that there is a trend towards higher risk (i.e. lower efficacy) by administered dose in the treatment group though it is not significant.

**Predicting efficacy of mAb against Omicron subvariants**

*Meta-analysis to identify IC-50's against Omicron subvariants:* In order to predict the efficacy of monoclonal antibody therapies to prevent severe outcomes in infections with the Omicron subvariants we required an estimate of the IC-50 of these mAbs against each variant. Fortunately, a systematic review published by Tao et al, provides a comprehensive analysis of reported IC-50s of various monoclonal antibodies against the ancestral SARS-CoV-2 virus as well as the BA.1 and BA.2 Omicron subvariants[18]. To supplement this systematic review with data on BA.4/5 we performed a search of the literature for studies that reported IC-50s of monoclonal antibodies against the BA.4 or BA.5 subvariants (which share a common spike amino-acid sequence[19]). Literature searches were performed in PubMed and Europe PMC with the search terms in **Table S16**. Search results to 14 July 2022 were included. Only primary research articles were included (no review articles, perspectives or editorials were included). After reviewing the abstracts of the search results, we identified four articles with the IC-50 of in vitro neutralisation reported for monoclonal antibodies against BA.4/5 (**Table S14**). Combining all the measured IC-50's and adopting the upper limit of detection of 10,000 ng/ml (which was the maximum mAb concentration used in the majority of assays in both the BA.4/5 studies as well as the BA.1 and BA.2 studies in the Tao et al systematic review), we performed a mixed effects linear regression with censoring of IC-50's when they were above the limit of detection of each assay and a random effect for the mean IC-50 (grouped by study) and fixed effects for each mAb IC-50 against each SARS-CoV-2 variant (using the *lmec* package in R[4,20]), to determine the mean IC-50 for each antibody against each variant, i.e.

$$IC50 \sim Variant * mAb + (1|Study).$$

Variant and mAb are categorical covariates. All antibodies included in a given study were included in the analysis, and only the ancestral, BA.1, BA.1.1, BA.2 and BA.4/5 variants were considered in the analysis. The estimated mean neutralisation IC-50s from this censored regression are shown in **Fig. S4** (black circles). Note that estimates of the mean that were above the limit of detection (i.e. 10,000 ng/ml) are unreliable and were removed from the figures.

<u>*Calibration of neutralisation IC-50s against Omicron subvariant to convalescent equivalence scale:*</u> The mean IC-50 for each antibody against each variant in **Fig. S4** was next converted to the 'fold-convalescent equivalence scale' (which is described in above section *Estimating the administered neutralising antibody dose relative to titres in convalescent individuals*). This was done by creating a calibration curve relating the IC-50s for each antibody against ancestral virus, from the meta-analysis (**Fig. S5**) with the IC-50s used to develop the convalescent equivalence scale (described above), which was based on a single study of *in vitro* neutralisation by McCallum et al[11] and which are shown in as blue circles in **Fig. S4**. Comparing the IC-50 for each mAb in the McCallum study with the mean reported in the meta-analysis, we see a strong correlation (**Fig. S5**). We perform a Deming regression (using a maximum likelihood method, with variance ratio assumed to be 1 between the x and y variables as both x and y are IC-50 measurements) to relate these two IC-50 scales and estimate the uncertainty in this relationship using the Hessian to compute the covariance matrix (**Fig. S5**). Using this calibration curve, we could relate the mean IC-50 for each antibody against each subvariant in **Fig. S4** to the IC-50 scale of the McCallum et al study. We then predicted the convalescent dose equivalent of individual monoclonal antibodies against the Omicron subvariant (as above, *Estimating the administered neutralising antibody dose relative to titres in convalescent individuals*).

<u>*Predicting therapeutic efficacy with confidence bands:*</u> Using the relationship between administered dose (fold-convalescent) of mAb and efficacy (**Fig. 2**) and equation 1 above, we predicted the expected efficacy of sotrovimab, Evusheld (cilgavimab+tixagevimab), bebtelovimab and cilgavimab alone (the component of Evusheld most active against BA.4/5) against the BA.1, BA.2 and BA.4/5 variants. We also predicted the effect of twice or four-times the dose of sotrovimab. The 95% Confidence bands on the model's predicted efficacy were estimated using parametric bootstrapping (as described in Cromer et al[21]). This boot-strapping procedure involved recomputing the estimated efficacy of each mAb and variant combination 100,000 times, sampling from distributions to account for the uncertainty (standard error) in the IC-50 of each mAb against the omicron subvariants from the meta-analysis (**Fig. S4**), the uncertainty (standard error) in the relationship between the meta-

analysis and the McCallum et al study (**Fig. S5**), and the uncertainty (covariance matrix of estimated parameters) in the model relating administered dose to efficacy (**Fig. 2**). The 95% CI was estimated using the percentile method. The p-value for whether the predicted efficacy was larger than 30% (**Fig. 3**) was computed by counting the proportion of boot-strapped estimated that were below 30% of the 100,000 estimates.

*Therapeutic efficacy in a non-naïve population*: To estimate the maximal efficacy of therapeutic passive Ab treatment in a non-naïve population, we assume that there exists a passive Ab treatment with the maximal efficacy of 70.2% for the given variant (**Table S8**), that the efficacy of antibodies for preventing progression to hospitalisation follows the same dose-response curve as passive antibody treatment (**Fig. 2**), and that the maximal efficacy that can be achieved is again 70.2%. The maximal efficacy of passive Ab treatment that would be estimated from a randomized control trial is then

$$\text{Efficacy} = 1 - \text{ relative risk} = 1 - \frac{\text{risk of treated patients}}{\text{risk of untreated patients}}.$$

The risk of treated patients is the fraction of treated patients who progressed from a symptomatic infection to hospitalisation. With a baseline risk *b* for all patients and passive Ab treatment with maximal effect *m*, the predicted fraction of treated patients progressing is $b \times (1-m)$. For untreated patients, the fraction of patients progressing is determined by the baseline risk and the level of protection they have due to their own antibodies, i.e. $b \times (1-E(\text{initial Abs}))$ where $E(.)$ denotes the dose-response curve for passive Ab treatment (**Fig. 2**). Overall, we thus find a maximal efficacy of passive Ab treatment in a non-naïve population of

$$\text{Efficacy} = 1 - \frac{\text{risk of treated patients}}{\text{risk of untreated patients}} = 1 - \frac{b \times (1-m)}{b \times \left(1 - E(\text{initial Abs})\right)} = \frac{m - E(\text{initial Abs})}{1 - E(\text{initial Abs})}.$$

This efficacy is shown in **Fig. 4** (blue curve).

We computed the 95% confidence bands for this efficacy using bootstrapping of the parameters of the efficacy function *E(.)* with 100,000 repetitions as described above (*Dose-response curve fitting* and *Predicting therapeutic efficacy with confidence bands*).

To estimate the maximal efficacy of passive Ab treatment in a non-naïve population against a variant, we used the same approach but rescaled the initial Ab titre to account for a drop in neutralisation titre against the variant. We assume again that there is some passive Ab treatment with maximal efficacy of 70.2%. Thus, for a variant with an *x*-fold drop in neutralisation titre against Ancestral, the estimated efficacy is

$$\text{Efficacy}_{\text{variant}} = \frac{m - E(\text{initial Abs } / \ x)}{1 - E(\text{initial Abs}/\ x)}.$$

We used a 19.2-fold drop of Omicron BA.4/5 against Ancestral as reported by Wang et al[22] to predict the maximal efficacy of passive Ab treatment in a non-naïve population against the Omicron BA.4 and 5 variants (**Fig. 4** black line and grey 95% confidence bands). As a reference, we add the neutralisation titre of patients who recently received 2 doses of an mRNA vaccine which is 3.25-fold early convalescent titre (mean of the fold-convalescent titre of Pfizer vaccinees (2.4) and Moderna vaccinees (4.1) as reported by Khoury et al[6]). By waning this titre will decrease over time and it may also be lower due e.g. immuno suppression but it may also by higher after boosting.

#### Supplementary results

##### Prophylactic passive antibody treatment to prevent infection

Interestingly, two studies of post-exposure prophylaxis administered to close contacts showed efficacies of 42.1% and 82.3%. One of these studies also reported an efficacy in preventing infection beyond one week after administration of antibodies of 92.8% (post-hoc analysis)[23], which we assume was effectively true prophylaxis (since exposure most likely occurred after treatment). Finally, one study investigating true prophylaxis showed an efficacy of 92.4% in preventing symptomatic SARS-CoV-2 infection (preprint publication[24]).

We have previously estimated that a mean level of neutralising antibody of around 20% of the convalescent titre provides 50% protection from acquisition of symptomatic SARS-CoV-2 infection (with the ancestral variant)[6]. Therefore, we first investigated whether the results of the two studies of prophylactic administration of monoclonal antibodies were consistent with this. **Fig. S7** shows the relationship between neutralisation and protection following infection or vaccination with different agents (data and model from Khoury et al and Cromer et al[6,15,21], in grey and black), and the range of antibody levels expected after boosting with an mRNA vaccine[21] (shaded blue). The administered dose of monoclonal antibody is shown on the horizontal axis (in fold-convalescent) and it is clear that this dose is significantly higher than the levels achieved even after vaccination and boosting (**Fig. S7**). However, the level of protection observed after high-dose passive antibody administration was similar to that seen after mRNA vaccination. This could be because the *in vivo* neutralising antibody titres were lower than might be predicted from the administered dose. Alternatively, this perhaps suggests a maximum level of protection from symptomatic SARS-CoV-2 infection that can be achieved by neutralising antibodies, which could not be estimated from the vaccine studies.

Previous studies have identified the antibody levels providing protection from COVID-19 after vaccination and suggest that a titre equivalent to 0.2-fold of the average early convalescent titre provides 50% protection from acquiring symptomatic SARS-CoV-2[6]. Here we estimate the level of antibody required to give 50% of the maximal protection from progression from symptomatic to severe infection and find an administered dose equivalent to 0.19-fold the mean convalescent titre. This is remarkably similar to the neutralisation level associated with protection from symptomatic infection in the vaccine studies and suggests that the major difference between prophylaxis and early therapeutic intervention is not in the level of antibody required, but rather the maximum protection that can be achieved.

##### Dose response for treatment of hospitalised individuals

As described above, studies of passive antibody administration to hospitalised patients showed low or no efficacy overall. This was the case even with administration of high doses of monoclonal antibodies, in a similar range of doses to those which were effective at earlier stages of infection (**Fig. S8**).

##### Using lower doses of mAbs can avert more hospitalisations when products are limited

To date, the availability of mAbs has been limited, and thus we explored the predicted hospitalisations that can be averted with lower doses of mAbs than those used in the studies so far. One strategy to allow treatment of more people is to give less compound per person, and this may be a viable strategy given that most studies of mAbs have used doses much higher than the EC-90 (**Fig. 2**, **Fig. S2** and **Table S12**). However, at lower doses the antibodies will be less effective for each treated individual. Thus, for each antibody included in the trials and analysed in this study, we analysed the lowest dose used and predicted the fold-increase in the hospitalisations averted by giving less of the compound per person and distributing the total dose amongst more individuals (see Supplementary methods *Fold increase of hospitalisations by administered mAb dose*). We find that decreasing the dose of each antibody to the EC-90 level, would avert between 7.1- and 710.1-fold more hospitalisations (**Fig. S9** and **Table S13**). Lower doses could avert even more cases with a maximum effect at a dose of 0.094-fold convalescent (less than 7mg for each considered mAb, **Table S13**). However, such a low dose is estimated to provide only 19.6% protection for treated individuals and thus would only be optimal when product availability is exceptionally low compared with the number of cases requiring treatment. Reducing administered doses below 0.094-fold of the mean convalescent titre equivalent allows distribution to more individuals but is predicted to result in more hospitalisations due to the low efficacy beginning to be a limiting factor.

#### Supplementary figures

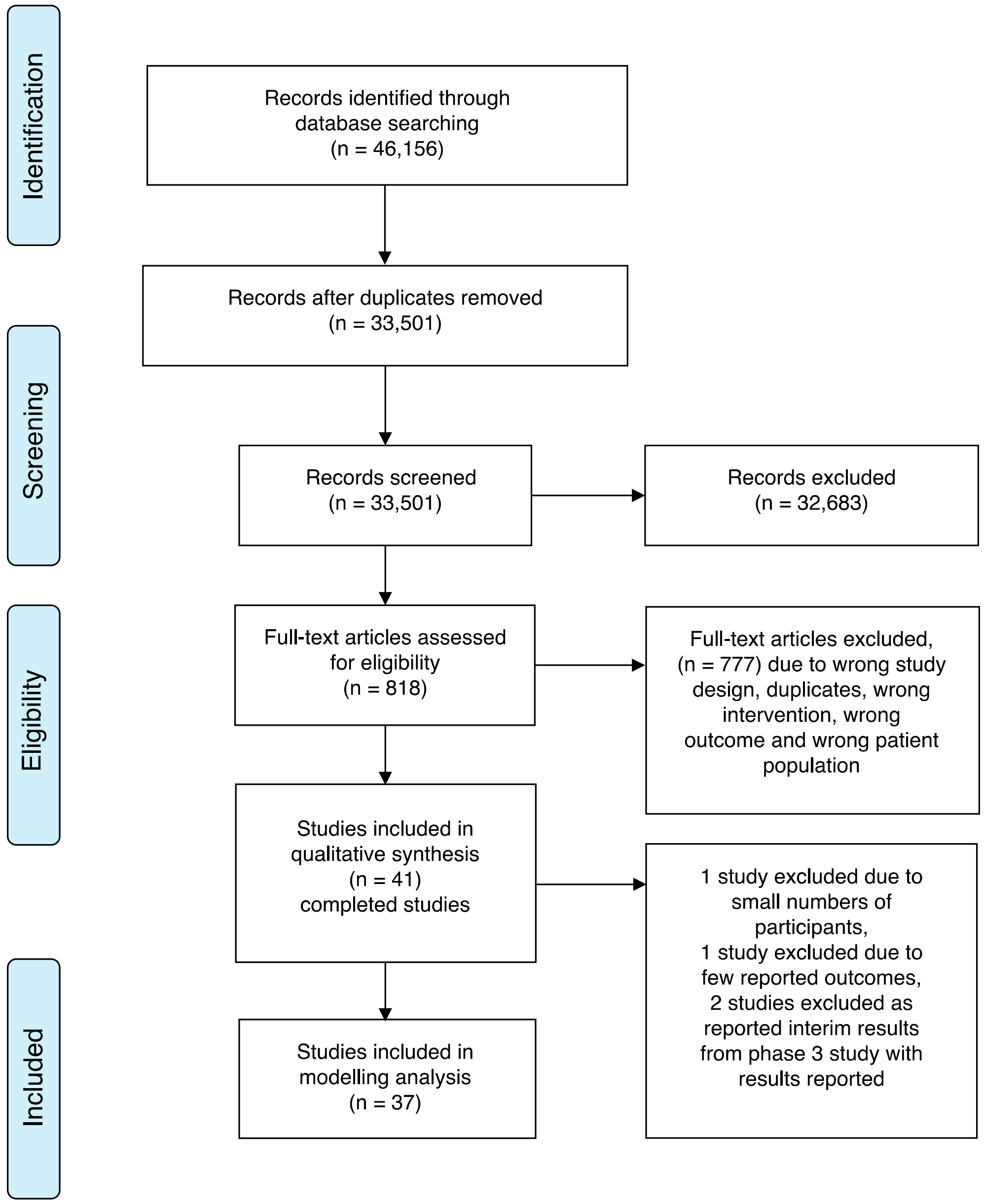


**Fig. S1** Studies searched, selected, and included in the analysis.

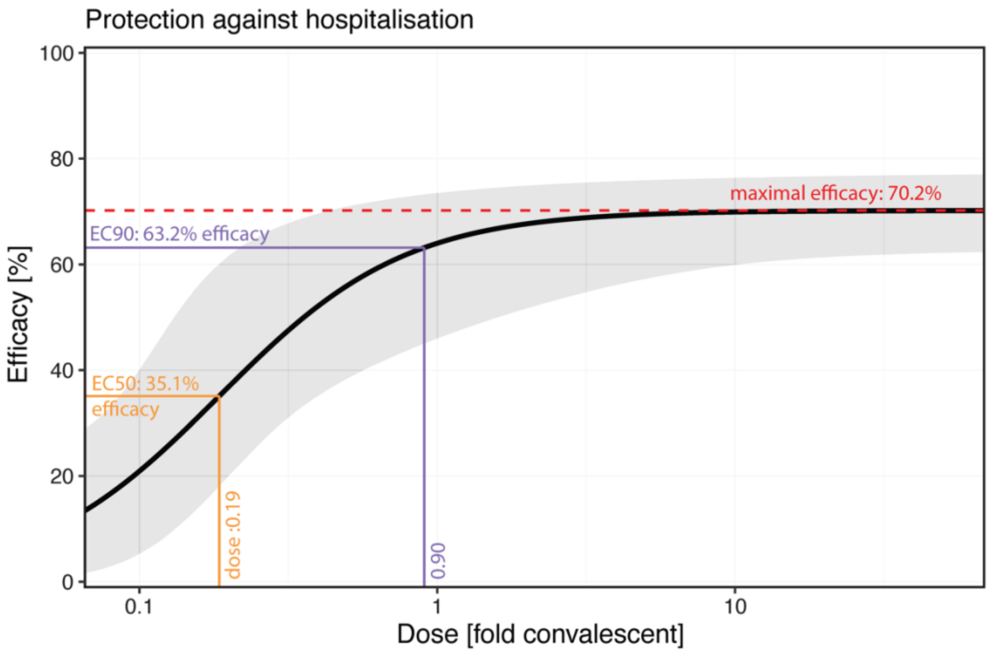


**Fig. S2** The dose-response curve for treatment of symptomatic ambulant subjects and efficacy in preventing progression to hospitalisation with maximal efficacy, EC-50, and EC-90 indicated.

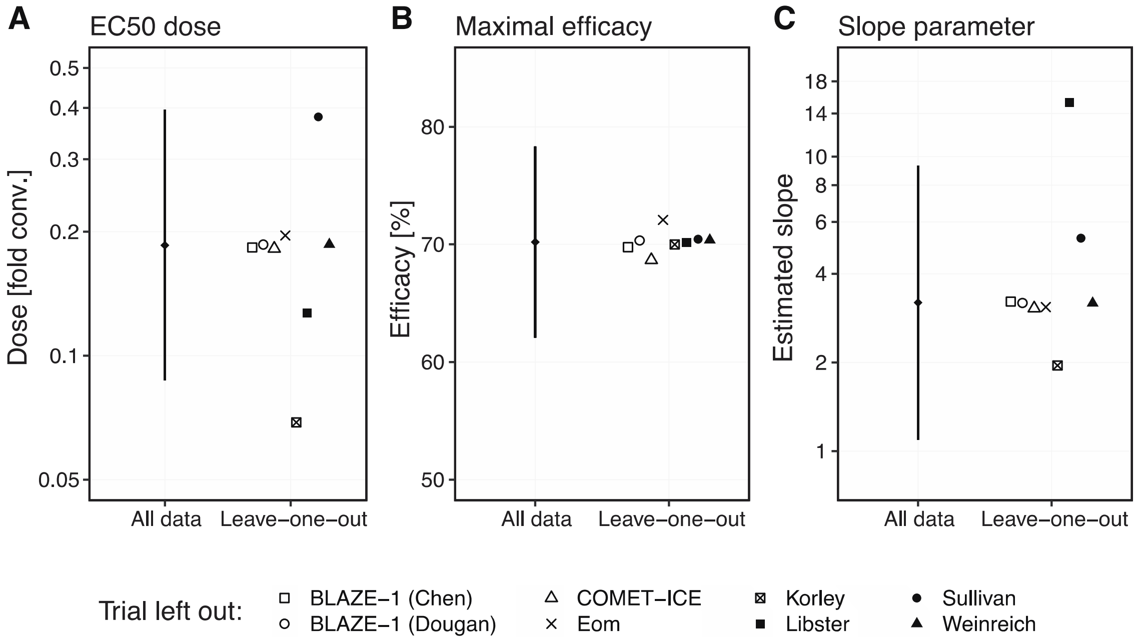


**Fig. S3** Leave-one-out analysis for the dose-response curve for preventing hospitalisation. We investigated the sensitivity of the three parameters for the logistic efficacy function, the EC50 dose (**A**), maximal efficacy (**B**), and slope parameter (**C**), by fitting the dose-response curve to the data excluding one study at a time. The parameter estimates with one omitted study agree well with the 95% CIs from the fit to all data, with only the EC-50 dose after omitting the Korley study[9] and the slope parameter after omitting the Libster study[8] as outliers.

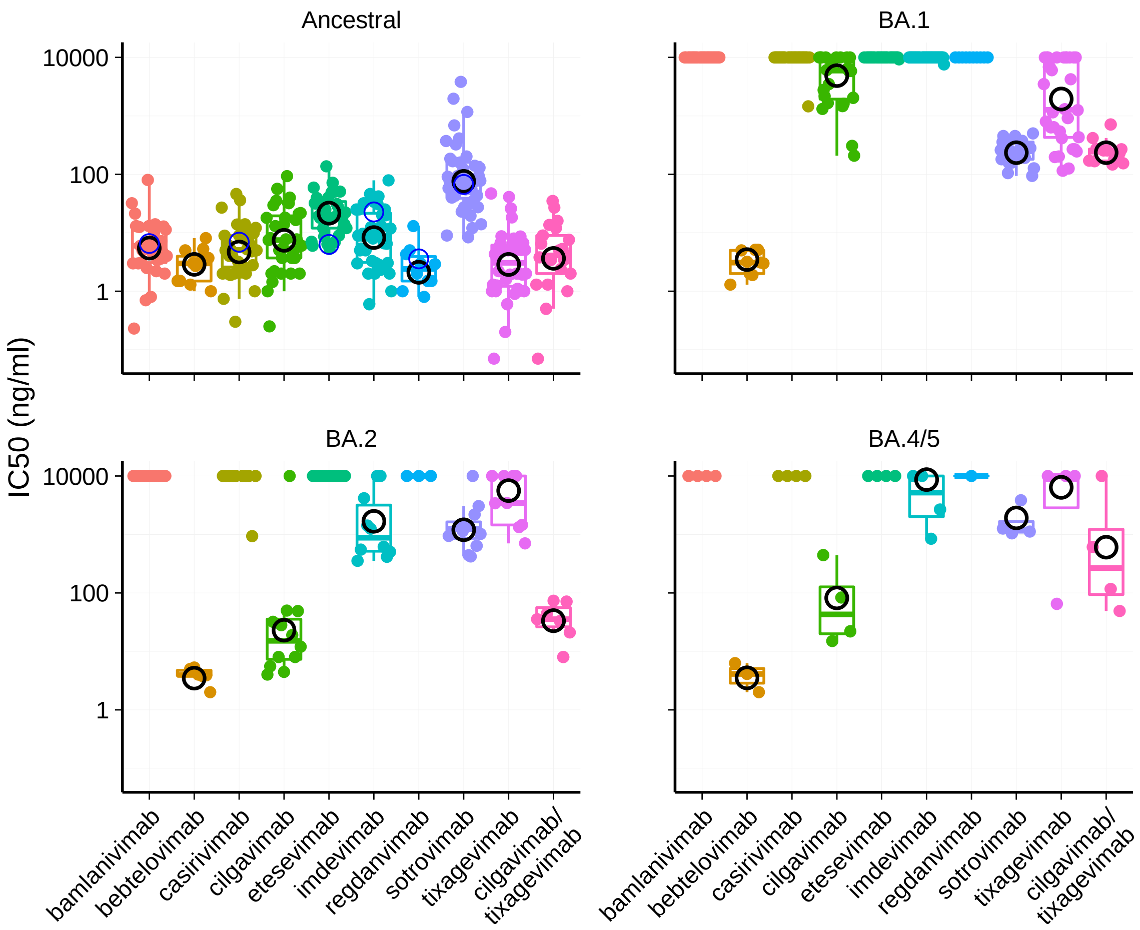


**Fig. S4** Estimated mean neutralisation IC-50s for different variants and mAbs. The black circles show the estimated mean neutralisation IC-50 using censored regression (see Supplementary methods). Estimates of the mean that were above the limit of detection (10,000 ng/mL) were removed from the figures as they are unreliable. The *in vitro* neutralisation by McCallum et al[11] against ancestral (which was used to compute the neutralisation on a 'fold convalescent' scale, see Supplementary methods) is indicated by blue circles.

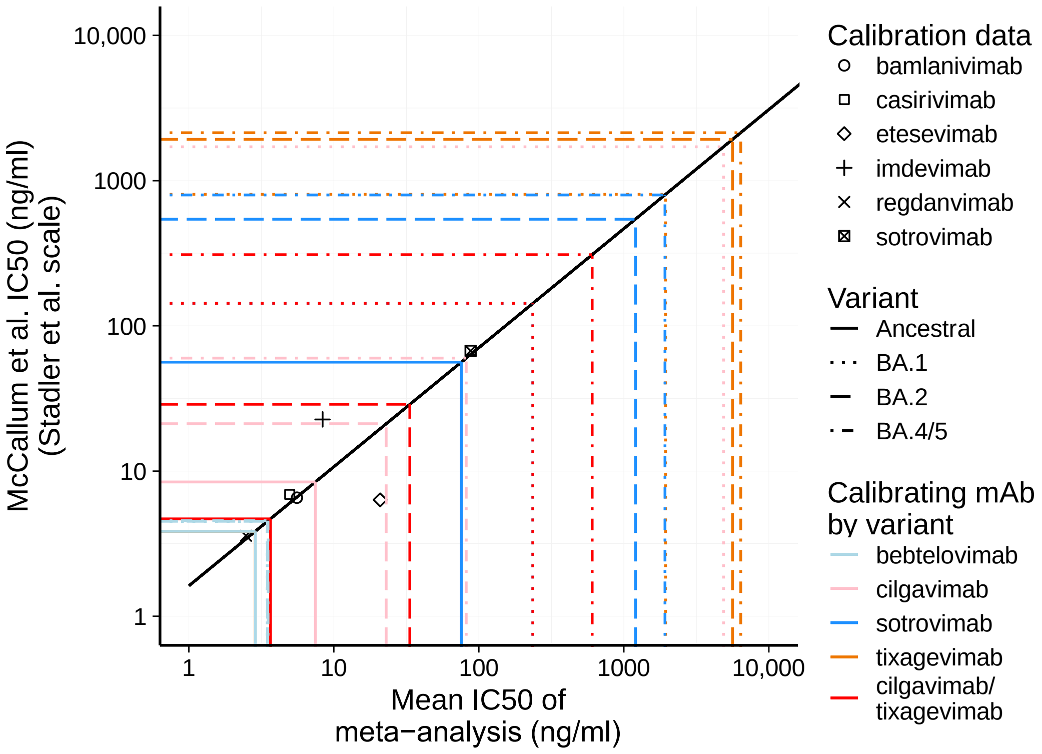


**Fig. S5** Calibration curve relating the IC-50s for each antibody against ancestral virus from the mean from the meta-analysis (horizontal axis) to the IC-50s in the McCallum et al study (vertical axis). Using Deming regression, we relate these two IC-50 scales and estimate the uncertainty (black line and 95% confidence bands in grey, details can be found in the Supplementary methods).

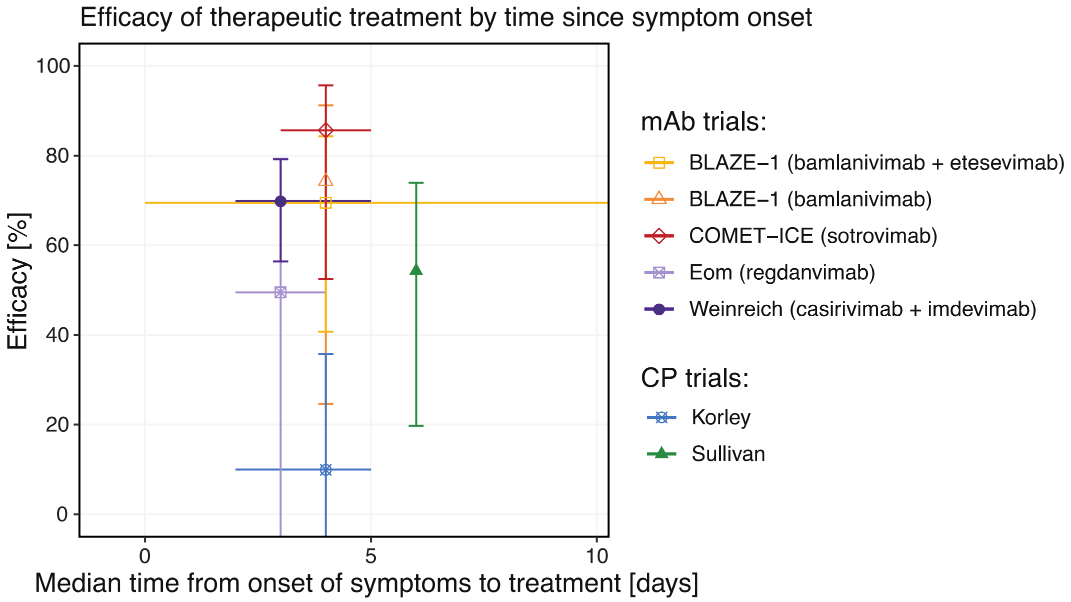


**Fig. S6** Efficacy of therapeutic treatment by time from symptom onset to treatment. For therapeutic treatment, i.e. treatment of symptomatic patients and prevention of hospitalisations, the efficacy of treatment is shown for different median times between symptom onset and treatment. Vertical bars denote 95% CIs for the efficacy of preventing hospitalisations and horizontal lines indicate the range of treatment days (for “BLAZE-1 (bamlanivimab + etesevimab)”, “COMET-ICE (sotrovimab)”, and “Sullivan”) or interquartile range of treatment days (for “Eom (regdanvimab)” and “Weinreich (casirivimab + imdevimab)”) timed from symptom onset. Note that “COMET-ICE (sotrovimab)” did not report a median time since symptom onset but a range of 3 to 5 days, we thus used a median of 4 days for visualisation.

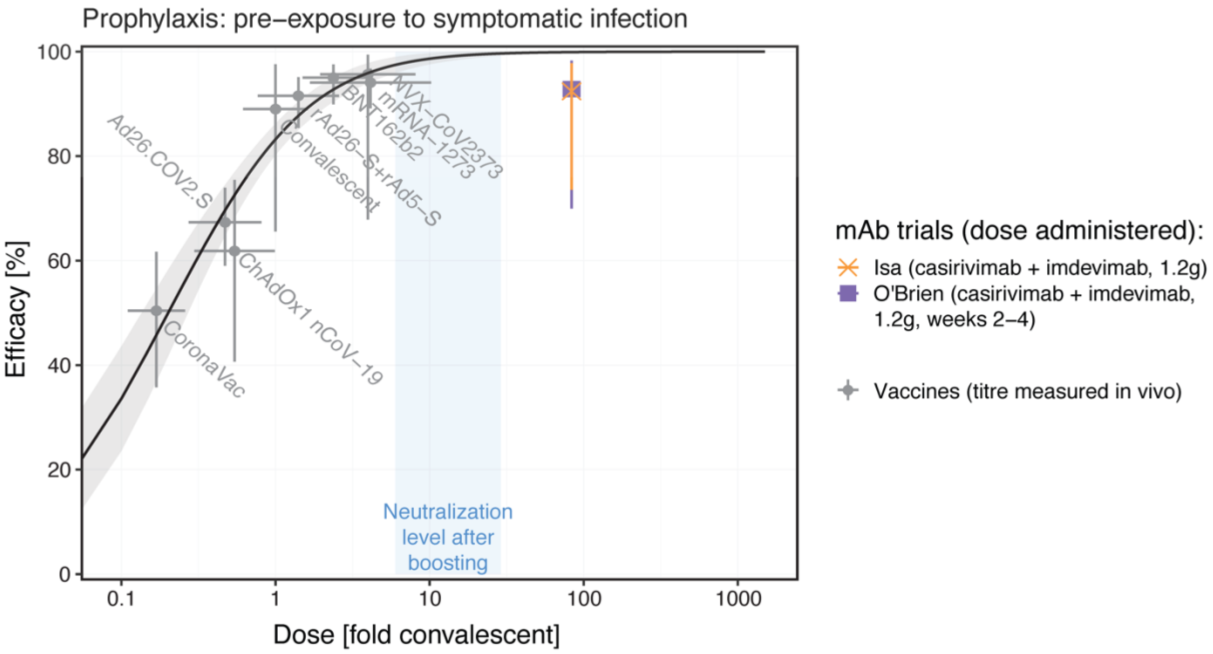


**Fig. S7** Estimating the dose-response curve for neutralising antibodies protecting from symptomatic SARS-CoV-2 infection. The curve and dots in grey show the relationship between neutralising antibody levels and protection from symptomatic SARS-CoV-2 infection previously determined in studies of vaccination[6]. The blue shaded area indicates the approximate neutralisation level of different studies of booster vaccination[21]. The purple and orange symbols and lines indicate the estimated administered dose and efficacy of monoclonal antibodies used in the prophylactic setting.

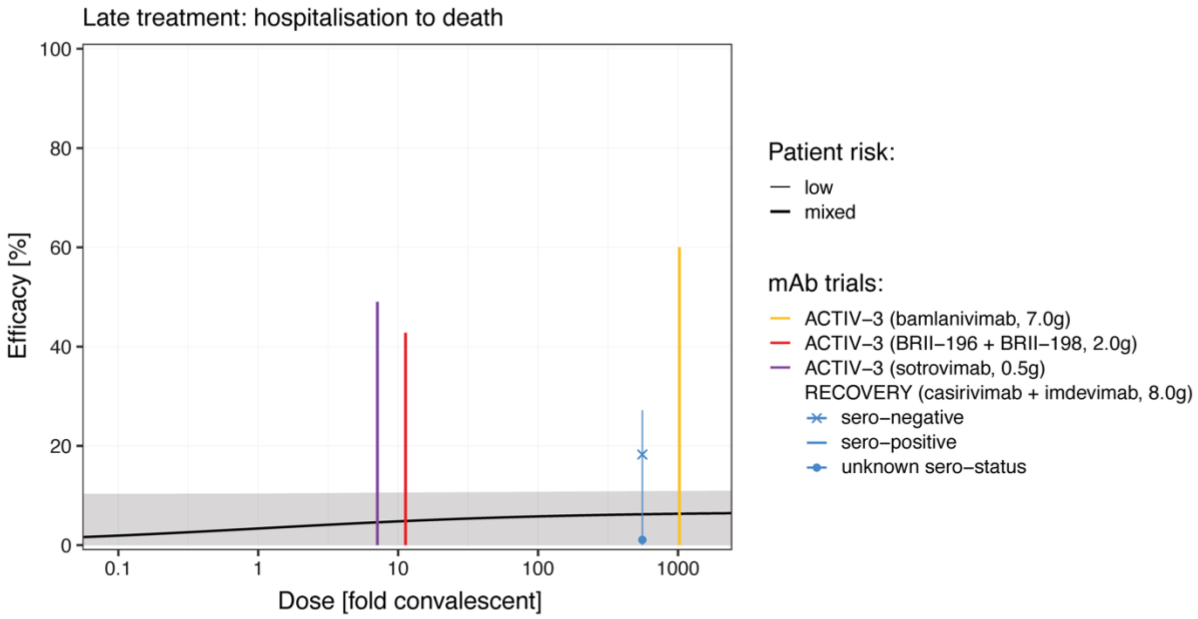


**Fig. S8** Dose-response curve for treatment of hospitalised subjects and prevention of death. No studies show significant efficacy for the treated population despite high antibody levels. Only the subgroup of sero-negative patients in the RECOVERY trial showed significant efficacy (18.3%, 95% CI: 8.3-27.2%). Apart from the sero-negative patients in the RECOVERY trial, only the patients with unknown sero-status at baseline in the RECOVERY had an estimated efficacy that was positive (1.1%, 95% CI: -19.7 to 18.3%). For all other subgroups and studies, efficacy was negative and thus only the 95% CI for efficacy is shown. The shaded area indicates the 95% confidence region for the fitted dose-response curve. For the estimated parameters for the dose-response curve, see **Table S15**.

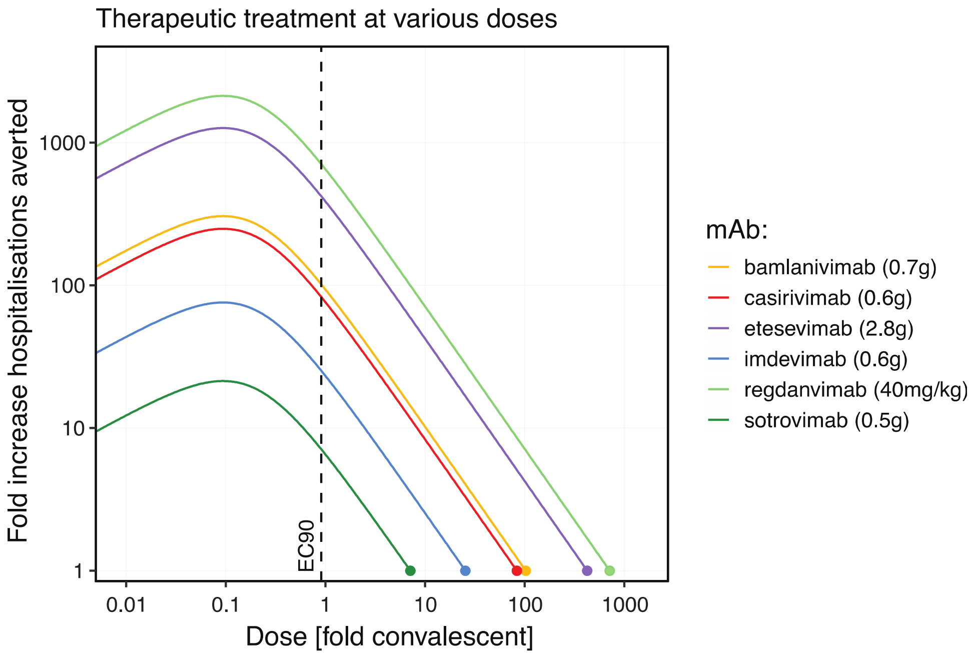


**Fig. S9** Predicted increase in hospitalisations averted for therapeutic treatment of ambulant symptomatic subjects with different doses of mAb. This figure shows the trade-off between treating more individuals at a lower dose and the reduced efficacy of treatment at a lower dose for various mAbs. Currently administered doses are indicated as a dot at '1-fold' hospitalisations averted. If more than one dose was used, only the lowest reported dose is shown here. The EC-90 dose is indicated by a vertical dashed black line. The maximal fold-increase in hospitalisations averted is achieved at a dose corresponding to 0.094-fold convalescent which would be a dose below 7mg for all shown mAbs (**Table S12**).

### Supplementary tables

**Table S1. Data sources for efficacy data for SARS-CoV-2 monoclonal antibody studies**

| Study | Number of partici-pants | Name of mAb | Dosage | Time of administration, median time from onset of symptoms to drug (days) | Population and disease stage | Measurement of effectiveness | Study Primary Outcome | Ref. | Data derived from |
|---|---|---|---|---|---|---|---|---|---|
| Isa | 969 | Casirivimab/ imdevimab | 1.2g (0.6g each) | NR | Uninfected: Pre-exposure | 1. Symptomatic infection | Incidence of AEs of special interest (AESIs), within 4 days of administration, concentrations of REGEN-COV in serum over time. | [24] | Figure 1 |
| O'Brien | 1505 | Casirivimab/ imdevimab | 1.2g | NR | Uninfected: Post- exposure (asymptomatic household contact with exposure to an individual with a diagnosis of SARS-CoV-2 infection, randomized within 96 hours of collection of the index cases' positive SARS-COV-2 diagnostic test sample) | 1. Infection<br>2. Symptomatic infection<br>3. Hospitalisation<br>4. All-cause mortality at 30 days | Proportion of individuals with symptomatic, RT-qPCR–confirmed SARS-CoV-2 infection during the 28-day EAP. | [25] | Table 2, S3, S9 in supplementary index of study reference |
| BLAZE-2 (Cohen) | 966 | Bamlanivimab | 4.2g | NR | Uninfected: Post-exposure (resident or staff in a skilled nursing or assisted living facility with at least one confirmed case of direct SARS-CoV-2 detection ≤7 days prior to randomization) | 1. Infection<br>2. Symptomatic infection<br>3. All-cause mortality reported at 57 days | Incidence of COVID-19, defined as the detection of SARS-CoV-2 by PCR and mild or worse disease severity within 21 days of detection, within 8 weeks of randomization. | [26] | Exploratory endpoints and eFigure 1, 3 of Supplementary index of study reference |
| Eom | 327 | Regdanvimab (CT-P59) | 40mg/kg, 80mg/kg | 3 | Infected: Ambulatory/mild | 1. Hospitalisation<br>2. ICU admission<br>3. Need for IMV<br>4. All-cause mortality at 30 days | Time to conversion to negative RT-qPCR result, time to clinical recovery | [27] | Table 1, 2, supplementary information |

| | | | | | | | | | |
|---|---|---|---|---|---|---|---|---|---|
| Weinreich | 5398 | Casirivimab/ imdevimab | 1.2g, 2.4g, 8.0g | 3 | Infected: Ambulatory/mild | 1. Hospitalisation 2. ICU admission 3. Need for IMV 4. All-cause mortality at 29 days | percentage of patients with ≥1 Covid-19-related hospitalisation or all-cause death through day 29 | [28] | Table 2, S7, S10, S12 |
| BLAZE-1 (Chen) | 467 | Bamlanivimab | 0.7g, 2.8g, 7.0g | 4 | Infected: Ambulatory/mild | 1. Hospitalisation 2. ICU admission 3. All-cause mortality at 30 days | Change in SARS-CoV-2 log viral load at day 11 | [29] | Table 3 |
| BLAZE-1 (Dougan) | 1035 | Bamlanivimab/ etesevimab | 2.8g/2.8g | 4 | Infected: Ambulatory/mild | 1. Hospitalisation 2. All-cause mortality at 30 days | COVID-related hospitalisation or death from any cause by day 29. | [30] | Figure 2 |
| COMET-ICE (Gupta) | 583 | Sotrovimab (VIR-7831) | 500mg | NR, range 3-5 | Infected: Ambulatory/mild | 1. Hospitalisation 2. ICU admission 3. Need for IMV 4. All-cause mortality at 30 days | Hospitalisation (for >24 hours) for any cause or death within 29 days | [31] | Table 2 |
| TACKLE | 822 | AZD7442, tixagevimab (AZD8895) and cilgavimab (AZD1061) | 0.6g, 2 doses | NR (within 5 days) | Infected: Ambulatory/mild | 1. All-cause mortality at 30 days | Composite of either severe COVID-19 or death from any cause through day 29 | [32] | Press release |
| ACTIV-3 | 314 | Bamlanivimab | 7.0g | 7 | Infected: Hospitalised/moderate to severe | 1. Need for IMV 2. All-cause mortality at 30 days | Sustained recovery during a 90-day period | [33] | Table 2 |
| ACTIV-3 (TICO) | 536 | Sotrovimab (VIR-7831), BRII-196 plus BRII-198 | 0.5g, 1g + 1g | 8 | Infected: Hospitalised/moderate | 1. Need for IMV 2. All-cause mortality up to 90 days | Time to sustained clinical recovery, defined as discharge from the hospital to home and remaining at home for 14 consecutive days, up to day 90 after randomisation. | [34] | Figure 3 |

| | | | | | | | | | |
|---|---|---|---|---|---|---|---|---|---|
| RECOVERY | 9785 | Casirivimab/ imdevimab | 8.0g | 9 | Infected: Hospitalised/ moderate to severe | 1. All-cause mortality at 30 days<br>2. Need for IMV reported as a composite outcome with death | 28-day mortality at seronegative participants and all participants | [35] | Table 2 |

**Table S2. Data sources for efficacy data for convalescent plasma and hyperimmune immunoglobulin studies**

| Study | Number of partici-pants | Dosage | Number of doses | Antibody levels | Time of administration, median time from onset of symptoms to drug (days) | Population and disease stage | Measurement of effectiveness | Study Primary Outcome | Ref. | Data derived from |
|---|---|---|---|---|---|---|---|---|---|---|
| Libster | 160 | 250ml | 1 | IgG titre against SARS-CoV-2 spike (S) protein (COVIDAR) > 1:1000 | NR (maximum 3) | Infected: Ambulatory/mild | 1. Hospitalisation<br>2. ICU admission<br>3. Need for IMV<br>4. All-cause mortality at 30 days | development of severe respiratory disease defined as a respiratory rate (RR) ≥ 30 and/or an O2 sat < 93% when breathing room air within 15 days | [8] | Table 2 |
| Korley | 511 | 250ml | 1 | SARS-CoV-2 pseudovirus reporter viral particle neutralization (RVPN) assay ≥1:250, or PRNT50 ≥1:250 | 4 | Infected: Ambulatory/mild | 1. Hospitalisation<br>2. Need for IMV<br>3. All-cause mortality | Number of patients with disease progression at 15 days | [9] | Table 1, 2, Figure 3 of study reference |
| Lopardo | 245 | 4mg/kg | 2 | NR | 6 | Infected: Hospitalised/moderate to severe | 1. ICU admission<br>2. Need for IMV<br>3. All-cause mortality at 30 days | Clinical improvement on ordinal scale at 28 days, hospital discharge | [36] | Table 2 |

| | | | | | | | | | | |
|---|---|---|---|---|---|---|---|---|---|---|
| Sullivan | 1181 | 250ml | 1 | EUROIMMUN IgG ELISA ≥ 1:320 | 6 | Infected: Ambulatory/mild (symptomatic outpatients) | 1. Hospitalisation<br>2. ICU admission<br>3. All-cause mortality at 30 days | Cumulative incidence of hospitalisation or death prior to hospitalisation at 28 days | [10] | Table 2 |
| CAPSID (Körper) | 105 | 250-325ml | 3 | Neutralising antibodies by PRNT50, median (1:160, IQR 1:80-1:320) | 7 | Infected: Hospitalised/moderate to severe | 1. All-cause mortality at 30 days | Composite endpoint of survival and no longer fulfilling criteria of severe disease at 30 days | [37] | Figure 2 of study reference, Figure 1A, 2A of supplementary table |
| Kirenga | 122 | 250-300ml | 2 | Acro Biosystems Anti-SARS CoV-2 antibody IgG titre Serological ELISA (Spike protein RBD), median 139.5 (IQR 84.3–195.4) AU | 7 | Infected: Ambulatory/mild | 1. All-cause mortality at 30 days | Time to viral clearance at 30 days | [38] | Table 1, 2, Figure 2, Supplementary Table 2 of study reference |
| Ortigoza | 941 | 250ml | 1 | Reactive on SARS-CoV-2 Microsphere Immunoassay, Ortho-Clinical Diagnostics VITROS signal to cut-off value ≥12 | 7 | Infected: Hospitalised/moderate to severe | 1. All-cause mortality at 30 days | Clinical status based on severity rating on WHO ordinal scale for clinical improvement at 14 days | [39] | Figure 2 |
| Agarwal | 1210 | 200ml | 2 | Microneutralization test, median 1:40 (IQR 1:30 to 1:80). Nab was tested at end of study: 63% of donors had Nab titre >1:20 with median titter 1:40 | 8 | Infected: Hospitalised/moderate to severe | 1. Need for IMV<br>2. All-cause mortality at 30 days | Composite of progression to severe disease (PaO2/ FiO2 <100 mm Hg) or all-cause mortality at 28 days | [40] | Table 1, 3 |

| | | | | | | | | | |
|---|---|---|---|---|---|---|---|---|---|
| ITAC | 593 | 0.4g/kg, capped at 40g | 1 | Sero-neutralization validated assay was calibrated against the WHO International standard for each lot of hIVIG | 8 | Infected: Hospitalised/moderate to severe | 1. All-cause mortality at 30 days | Clinical status by seven-category ordinal endpoint at 7 days | 41 | Table 2 |
| Simonovich | 228 | 415-600ml | 1 | IgG titre against SARS-CoV-2 spike (S) protein (COVIDAR), median 1:3200 (IQR 1:800-1:3200) | 8 | Infected: Hospitalised/moderate | 1. ICU admission<br>2. Need for IMV<br>3. All-cause mortality at 30 days | Clinical status based on severity rating on WHO ordinal scale for clinical improvement at 30 days | 42 | Table 1 |
| CONCOR1 (Bégin) | 921 | 500ml (or 250ml) | 1 (or 2) | 4 assays used including ADCC ratio, anti RBC ELISA anti S IgG, PRNT50. Each had different criteria that were based on the presence of either viral neutralizing antibodies at a titre of >1:160 or antibodies against the receptor binding domain (RBD) of the SARS-CoV-2 Spike protein at a titre of >1:100 | 8 | Infected: Hospitalised/moderate to severe (excluding intubated patients) | 1. All-cause mortality at 30 days | Need for intubation or patient death at 30 days | 43 | Figure 2 of study reference |
| Bennett-Guerrero | 74 | 240ml | 2 | NT50 were median 1:334 (IQR 1:192–1:714) in a pseudotype assay and median 1:526 (IQR 1:359–1:786) in a plaque neutralization assay (PRNT) (gold standard) | 9 | Infected: Hospitalised/moderate to severe | 1. All-cause mortality at 30 days[1] | number of days patient remained ventilator-free at 28 days | 44 | Figure 3 of study reference |
| RECOVERY (Horby) | 11558 | 200-350ml | 2 | EUROIMMUN IgG enzyme-linked immunosorbent assay (ELISA) test targeting the spike (S) glycoprotein, sample to cut-off ratio of ≥6.0 | 9 | Infected: Hospitalised/moderate to severe | 1. Need for IMV<br>2. All-cause mortality at 30 days | All-cause mortality at 28 days | 45 | Table 2, Webtable 1, Webfigure 1, 4 in Supplementary Index |

| | | | | | | | | | | |
|---|---|---|---|---|---|---|---|---|---|---|
| O'Donnell | | 250ml | 1 | anti-SARS-CoV-2 total IgG antibody titre by quantitative ELISA. Neutralization titre was also determined with a SARS-CoV-2 viral neutralization assay which measured inhibition of virus growth after exposure to serial plasma dilutions using qRT-PCR, median 1:160 (IQR 1:80 to 1:320) | 9 | Infected: Hospitalised/moderate to severe | 1. All-cause mortality at 30 days[2] | Clinical status at 28 days following randomization using 7-point ordinal scale | [46] | Table 2 |
| PLACOVID (Sekine) | 160 | 300ml | 10 | Neutralising antibody via cytopathic effect-based virus neutralization test (CPE-based VNT), median 1:320 (IQR 1:160 to 1:960) | 10 | Infected: Hospitalised/moderate to severe | 1. Need for IMV<br>2. All-cause mortality at 30 days | Clinical improvement (improvement of 2 points from randomisation in a 6-point ordinal severity scale) at 28 days | [47] | Table 2 of study reference |
| Li | 103 | 200-300ml | 1 | S-RBD–specific IgG titre ≥ 1:640 correlating to serum neutralisation titre of 1:80 | 30 | Infected: Hospitalised/moderate to severe | 1. All-cause mortality at 30 days | Clinical improvement (patient discharged alive or reduction of 2 points on a 6-point disease severity scale) within 28 days | [48] | Table 3 |
| Ali | 50 | 4 arms: 0.15g/kg, 0.20g/kg, 0.25g/kg, 0.30g/kg | 1 | Plasma tested, with a lower limit of 10 cut-off index (COI), measurement through electrochemiluminescence immunoassay analyzer (ECLIA) | NR | Infected: Hospitalised/moderate to severe | 1. All-cause mortality at 30 days | Mortality at 28 days, Clinical status on ordinal scale, Horowitz index | [49] | Table 2 |

| | | | | | | | | | |
|---|---|---|---|---|---|---|---|---|---|
| AlQahtani | 40 | 200ml | 2 | NR | NR | Infected: Hospitalised/moderate to severe | 1. Need for IMV<br>2. All-cause mortality at 30 days | Requirement for ventilation (non-invasive or mechanical) | [50] | Table 3 |
| Bajpai | 51 | 250ml | 2 | S-RBD–specific IgG titre, median ≥640 (IQR 10, ≥640), SARS-CoV-2 Surrogate Virus Neutralization Test (sVNT) Kit (Genscript) median ≥80 (IQR 10, ≥80) | NR | Infected: Hospitalised/moderate to severe | 1. Need for IMV<br>3. All-cause mortality at 30 days | Proportion of participants remaining free of mechanical ventilation at 7 days | [51] | Table 2 |
| Beltran Gonzalez | 190 | 200ml | 2 | Presence of anti-SARS-CoV-2 IgG by immunochemiluminescence (Architect Abbott) | NR | Infected: Hospitalised/severe | 1. All-cause mortality at 30 days[2] | Mean hospitalisation time | [52] | Study text under “Outcomes” |
| Menichetti | 487 | 200ml | 1-3 | Microneutralization test ≥1:160 | NR | Infected: Hospitalised/moderate to severe | 1. All-cause mortality at 30 days | composite of worsening respiratory failure ($PaO_2/FiO_2$ ratio <150 mm Hg) or death within 30 days | [53] | eTable 7 and eFigure 2 in Supplement 2 |
| Parikh | 60 | 30ml | 2 | NR | NR | Infected: Hospitalised/moderate to severe | 1. All-cause mortality at 30 days | mean change from day 1 to day 8 in an 8-point ordinal scale | [54] | Figure 1 |
| Pouladzadeh | 62 | 500ml | 1 | Strong positive results of the SARS-CoV-2 IgG/IgM Quick Test (German) for neutralizing IgG antibodies and negative results for IgM antibodies | NR (minimum 7) | Infected: Hospitalised/moderate to severe | 1. Need for IMV<br>2. All-cause mortality at 30 days | Improvement in the levels of cytokine storm indices | [55] | Table 1 |
| Ray | 80 | 200ml | 2 | EUROIMMUN IgG enzyme-linked immunosorbent assay (ELISA) test targeting the spike (S) glycoprotein, ≥1.5 for the ratio optical density between the sample and calibrator. | NR (5-10 days from initial presentation) | Infected: Hospitalised/moderate to severe (excluding intubated patients) | 1. Need for IMV<br>2. All-cause mortality at 30 days | All-cause mortality at 30 days | [56] | Figure 4 |

| | | | | | | | | | | |
|---|---|---|---|---|---|---|---|---|---|---|
| REMAP CAP (Estcourt) | 2011 | 200-350ml | 2 | Variable assays used by country, including EUROIMMUN IgG ELISA test sample to cut-off ratio of ≥6.0, virus microneutralization assay using Vero E6 cell ≥1:80, Abbott Architect SARS-CoV-2 IgG CMIA, PRNT50 ≥1:160, antibody-dependent cell cytotoxicity (ADCC), ELISA test targeting the spike (S) glycoprotein. | NR (within 48 hours of randomisation) | Infected: Hospitalised/moderate to severe | 1. All-cause mortality at 30 days | All-cause mortality at 90 days, Days alive and not receiving organ support in ICU | [57] | Figure 2,3 |

[1]In this study, note that the comparator arm was administered IVIG (not placebo)

[2]In this study, note that the comparator arm was administered standard plasma (not placebo)

**Table S3. Efficacy for preventing different stage progressions for pooled data**

| Stage transition | Efficacy in percent (95% CI) | |
|---|---|---|
| | mAb treatment | CP or hIVIG treatment |
| pre-exposure to symptomatic | 91.9 (81.3 – 97.2) | - |
| peri-(post-)exposure to symptomatic | 52.7 (32.6 – 67.3) | - |
| symptomatic to hospitalisation | 70.0 (60.5 – 77.5) | 31.4 (11.3 – 47.2) |
| symptomatic to ventilation | 71.2 (5.9 – 93.5) | 44.8 (-58.1 – 82.9) |
| symptomatic to death | 62.5 (39.0 – 77.8) | -5.3 (-104.8 – 45.64) |
| hospitalisation to ventilation | -51.7 (-230.7 – 27.4) | 1.9 (-7.6 – 10.6) |
| hospitalisation to death | 5.4 (-2.2 – 12.5) | 2.7 (-3.0 – 8.1) |

**Table S4. The effect of the treatment stage on efficacy for mAb treatment**

| *Progression to the next stage*[†] | | |
|---|---|---|
| **Variable** | **Relative risk (95% CI)** | **$\chi^2$ test p-value** |
| treatment | 0.12 (0.08 – 0.19) | $2.80 \times 10^{-27}$ |
| initial stage (numerical) | 1.20 (0.89 – 1.61) | - |
| treatment : initial stage (numerical) | 1.96 (1.70 – 2.28) | $6.11 \times 10^{-23}$ |

| *By outcome stage* | | | |
|---|---|---|---|
| **Outcome** | **Initial stage** | **Relative risk (95% CI)** | **$\chi^2$ test p-value** |
| symptomatic | peri-(post-)exposure (relative to pre-exposure) | 6.26 (2.48 – 19.21) | $3.57 \times 10^{-5}$ |
| ventilation | hospitalisation (relative to symptomatic) | 6.08 (1.25 – 45.39) | $2.40 \times 10^{-2}$ |
| death | hospitalisation (relative to symptomatic) | 2.00 (1.20 – 3.45) | $7.17 \times 10^{-3}$ |

[†] We consider the following stage transition only (to avoid a higher influence of studies that reported more outcomes): pre-exposure to symptomatic, peri-(post-)exposure to symptomatic, symptomatic to hospitalisation, and hospitalisation to death. All initial stages were transformed to numerical; pre-exposure was transformed to 0, peri-(post-)exposure to 1, symptomatic to 2, and hospitalisation to 3.

**Table S5. The effect of the treatment stage on efficacy for CP and hIVIG treatment**

| *Progression to the next stage*[†] | | |
|---|---|---|
| **Variable** | **Relative risk (95% CI)** | **$\chi^2$ test p-value** |
| treatment | 0.24 (0.08 – 0.68) | 0.0071 |
| initial stage (numerical) | 1.03 (0.49 – 2.15) | - |
| treatment : initial stage (numerical) | 1.42 (1.09 – 1.86) | 0.0089 |

| *By outcome stage* | | | |
|---|---|---|---|
| **Outcome** | **Initial stage** | **Relative risk (95% CI)** | **$\chi^2$ test p-value** |
| ventilation | hospitalisation (relative to symptomatic) | 1.78 (0.62 – 5.75) | 0.290 |
| death | hospitalisation (relative to symptomatic) | 0.76 (0.37 – 1.53) | 0.445 |

[†] We consider the following stage transition only (to avoid a higher influence of studies that reported more outcomes): pre-exposure to symptomatic, peri-(post-)exposure to symptomatic, symptomatic to hospitalisation, and hospitalisation to death. All initial stages were transformed to numerical; pre-exposure was transformed to 0, peri-(post-)exposure to 1, symptomatic to 2, and hospitalisation to 3.

**Table S6. Number of different outcome events for symptomatic patients across all trials**

| | mAb treatment | | CP treatment | |
|---|---|---|---|---|
| **Outcome** | **Treatment group (n)** | **Control group (n)** | **Treatment group (n)** | **Control group (n)** |
| hospitalisation | 64 (4038) | 193 (3737) | 80 (927) | 118 (923) |
| ventilation | 3 (2586) | 10 (2484) | 5 (330) | 9 (328) |
| death | 20 (4445) | 54 (4165) | 17 (998) | 16 (990) |

**Table S7. Dose fold-convalescent for mAb and CP trials**

| Trial | Treatment | Dose/Volume | Dose [fold convalescent] |
|---|---|---|---|
| BLAZE-1 (Chen) | mAb | bamlanivimab (0.7g)<br>bamlanivimab (2.8g)<br>bamlanivimab (7.0g) | 102.37<br>409.92<br>1,023.71 |
| BLAZE-1 (Dougan) | mAb | bamlanivimab + etesevimab (2.8g +2.8g) | 424.37 |
| COMET-ICE | mAb | sotrovimab (0.5g) | 7.11 |
| Eom | mAb | regdanvimab (40mg/kg)<br>regdanvimab (80mg/kg) | 713.41<br>1,427.61 |
| Isa | mAb | casirivimab + imdevimab (1.2g) | 83.45 |
| O'Brien | mAb | casirivimab + imdevimab (1.2g) | 83.45 |
| Weinreich | mAb | casirivimab + imdevimab (1.2g)<br>casirivimab + imdevimab (2.4g)<br>casirivimab + imdevimab (8.0g) | 83.45<br>166.90<br>556.19 |
| Korley | CP | 250mL | 0.097 |
| Libster | CP | 250mL (CP 1,000 to 3,200)<br>250mL (CP >3,200) | 0.38<br>1.47 |
| Sullivan | CP | 250mL | 0.153 |

**Table S8. Estimated parameters for the dose-response curve for preventing hospitalisations**

| Description | Estimate | 95% CI |
|---|---|---|
| Maximal efficacy | 70.2% | 62.1 – 78.3% |
| EC-50 dose (dose for half-maximal efficacy) | 0.185-fold conv. | 0.087 – 0.395-fold conv. |
| EC-90 dose (dose for 90% of the maximal efficacy) | 0.904-fold conv. | 0.208 – 17.803-fold conv. |
| IC50 dose (dose for 50% efficacy) | 0.357-fold conv. | 0.132 – 1.598-fold conv. |
| Slope parameter of the dose-response curve | 3.193 | 1.093 – 9.328 |

**Table S9. Efficacy of mAb treatment for preventing hospitalisation by dose**

| | Relative Risk (95% CI) | $\chi^2$ test p-value |
|---|---|---|
| treatment | 0.082 (0.018 – 0.315) | 0.00013 |
| $\log_{10}$(dose) | 0.998 (0.699 – 1.408) | - |
| mixed risk | 0.651 (0.345 – 1.217) | 0.17 |
| high risk | 0.567 (0.316 – 1.028) | |
| $\log_{10}$(dose) : treatment | 1.720 (0.991 – 3.155) | 0.054 |

**Table S10. Predicted therapeutic efficacy of mAbs against Omicron variants BA.1, BA.2 and BA.4/5**

| mAb | Dose [mg] | Efficacy [%] (95% CI) against Omicron variants | | |
|---|---|---|---|---|
| | | BA.1 | BA.2 | BA.4/5 |
| cilgavimab/tixagevimab | 600 | 69.2 (51.7 – 75.7) | 70.1 (61.1 – 76.6) | 67.4 (33.8 – 74.9) |
| cilgavimab | 300 | 32.7 (0.3 – 70.8) | 70.0 (60.4 – 76.5) | 69.4 (53.9 – 75.8) |
| tixagevimab | 300 | 50.0 (3.4 – 72.3) | 29.9 (0.2 – 70.7) | 27.4 (0.1 – 70.6) |
| bebtelovimab | 175 | 70.1 (61.9 – 76.8) | 70.1 (61.9 – 76.8) | 70.1 (61.8 – 76.8) |
| sotrovimab | 500 | 69.0 (50.5 - 75.5) | 63.0 (16.9 – 73.9) | 58.7 (6.1 – 73.4) |
| | 1000 | 69.7 (56.0 – 76.0) | 67.2 (31.4 – 74.9) | 65.3 (16.2 – 74.5) |
| | 2000 | 69.7 (59.5 – 76.4) | 67.2 (43.9 – 75.6) | 65.3 (30.6 – 75.3) |

**Table S11. Dose fold-convalescent for various mAb treatments**

| Monoclonal Ab | Dosage | mAb conc. [mg/mL] | IC-50 [ng/mL] | Fold IC-50 | Dose fold-convalescent |
|---|---|---|---|---|---|
| bamlanivimab (LY-CoV555, LY3819253) | 0.7g | 0.233 | 6.548 | 35,583.38 | 102.37 |
| | 2.8g | 0.933 | | 142,486.3 | 409.92 |
| | 7.0g | 2.333 | | 355,833.8 | 1,023.71 |
| casirivimab | 0.6g | 0.2 | 6.895 | 29,006.53 | 83.45 |
| | 1.2g | 0.4 | | 58,013.05 | 166.9 |
| | 4.0g | 1.333 | | 193,328.5 | 556.19 |
| etesevimab | 2.8g | 0.933 | 6.325 | 147,509.9 | 424.37 |
| imdevimab | 0.6g | 0.2 | 22.658 | 8,826.90 | 25.39 |
| | 1.2g | 0.4 | | 17,653.81 | 50.79 |
| | 4.0g | 1.333 | | 58,831.32 | 169.25 |
| regdanvimab (CT-P59) | 40mg/kg | 0.888 | 3.581 | 247,975.4 | 713.41 |
| | 80mg/kg | 1.777 | | 496,230.1 | 1,427.61 |
| sotrovimab (VIR-7831) | 0.5g | 0.166 | 67.125 | 2,472.998 | 7.11 |

**Table S12. mAb dose for various efficacies of preventing hospitalisation**

| Monoclonal Ab | EC-50 [mg] (35.1%) | EC-90 [mg] (63.18%) | Doses given [mg] | Fold-difference between administered doses and EC-90 dose |
|---|---|---|---|---|
| bamlanivimab | 1.27 | 6.17 | 700, 2800, or 7000 | 113.5, 453.8, or 1134.5 |
| casirivimab | 1.33 | 6.50 | 600, 1200, or 4000 | 92.3, 184.6, or 615.4 |
| etesevimab | 1.22 | 5.96 | 2800 | 469.8 |
| imdevimab | 4.38 | 21.36 | 600, 1200, or 4000 | 28.1, 56.2, or 187.3 |
| regdanvimab | 0.69 | 3.38 | 3200 or 6400* | 946.7 or 1893.5 |
| sotrovimab | 12.98 | 63.29 | 500 | 7.9 |

* Dose of 40 or 80mg/kg for an 80kg person.

**Table S13. Fold increase in hospitalisations averted**

| Monoclonal Ab | Dose given [mg] | EC-90 dose [mg] (63.18%) | Fold-increase in hospitalisations averted with EC-90 dose | Maximal fold-increase in hosp. averted | Dose [mg] for maximal fold-increase in hosp. averted |
|---|---|---|---|---|---|
| bamlanivimab | 700 | 6.17 | 101.9 | 305.5 | 0.64 |
| casirivimab | 600 | 6.50 | 83.1 | 249.0 | 0.67 |
| etesevimab | 2800 | 5.96 | 422.4 | 1,266.2 | 0.62 |
| imdevimab | 600 | 21.36 | 25.3 | 75.8 | 2.21 |
| regdanvimab | 3200 | 3.38 | 710.1 | 2,128.6 | 0.35 |
| sotrovimab | 500 | 63.29 | 7.1 | 21.3 | 6.56 |

**Table S14. Studies reporting the IC-50 from in vitro neutralisation experiments for monoclonal antibodies against Omicron subvariant BA.4/5**

| Study | Neutralisation assay | Variants tested | Monoclonal antibodies tested | Ref |
|---|---|---|---|---|
| Arora et al. | Pseudoviral neutralisations assay | B.1, BA.1, BA.2, BA.4/5 (also BA.2.12.1) | Casrivimab, Imdevimab, Bamlanivimab, Etesevimab, Cilgavimab, Tixagevimab, Regdanvimab, Bebtelovimab, Sotrovimab, Casirivimab plus Imdevimab, Bamlanivimab plus Etesevimab, Cilgavimab plus Tixagevimab (also S2H97) | 58 |
| Tuekprakhon et al. | Pseudoviral neutralisations assay | Victoria, BA.1, BA.2, BA.4/5 (also BA.1.1 and BA.3) | Imdevimab, Casirivimab, Cilgavimab, Tixagevimab, Bamlanivimab, Etesevimab, Sotrovimab, Cilgavimab plus tixagevimab (also ADG10, ADG20, ADG30) | 59 |
| Wang et al. | Pseudoviral neutralisations assay | D614G, BA.1, BA.2, BA.4/5 (also BA.1.1, BA.2.12.1) | Etesevimab, Casirivimab, Tixagevimab, Bamlanivimab, Imdevimab, Cilgavimab, Bebtelovimab, Sotrovimab, Casirivimab plus Imdevimab, Cilgavimab plus Tixagevimab, Bamlanivimabplus Etesevimab (also amubarvimab, romlusevimab, ADG-2, DH1047, S2X259, CAB-A17, ZCB11) | 22 |
| Yamasoba et al. | Pseudoviral neutralisations assay. | B.1.1, BA.2, BA.4/5 (also BA.2.11, BA.2.12.1, BA.2 L452Q, BA.2 S704L, BA.2 HV69-70del, BA.2 F486V, BA.2 R493Q) | Bamlanivimab, Bebtelovimab, Casirivimab, Cilgavimab, Etesevimab, Imdevimab, Sotrovimab, Tixagevimab, Casirivimab plus imdevimab, Etesevimab plus bamlanivimab, Cilgavimab plus tixagevimab | 60 |

**Table S15. Estimated parameters for the dose-response curve for treatment of hospitalised subjects preventing death**

| Description | Estimate | 95% CI |
|---|---|---|
| Maximal efficacy | 6.74% | -8.15 – 21.62% |
| EC-50 dose (dose for half-maximal efficacy) | 1.016-fold conv. | 0.494 – 2.081-fold conv. |
| Slope parameter of the dose-response curve | 0.910 | 0.0035 – 237.977 |

**Table S16. Search to identify studies of Omicron subvariant BA.4/5**

| Database | Date range | Search terms |
|---|---|---|
| PubMed | 1 March to 14 July 2022 | (omicron[ti] OR BA4[TI] OR BA5[TI] OR BA.4[TI] OR BA.5[TI] OR BA4/5[TI] OR BA.4/5[TI]) AND (mabs[ti] OR antibod*[ti] OR neutral*[ti] OR vitro[TI] OR in-vitro[TI] OR sotrovimab[ti] OR casirivimab[ti] OR imdevimab[ti] OR tixagevimab[ti] OR cilgavimab[ti]) AND 2022/03:3000[edat] |
| Europe PMC | 1 January to 14 July 2022 | ((TITLE:"omicron" OR (TITLE:"BA4") OR (TITLE:"BA5") OR (TITLE:"BA.4") OR (TITLE:"BA.5") OR (TITLE:"BA4/5") OR (TITLE:"BA.4/5")) AND ((TITLE:"mabs") OR (TITLE:"antibody") OR (TITLE:"antibodies") OR (TITLE:"neutralising") OR (TITLE:"neutralizing") OR (TITLE:"neutralisation") OR (TITLE:"neutralization") OR (TITLE:"vitro") OR (TITLE:"in-vitro") OR (TITLE:"sotrovimab") OR (TITLE:"casirivimab") OR (TITLE:"imdevimab") OR (TITLE:"tixagevimab") OR (TITLE:"cilgavimab")) AND (SRC:PPR)) AND (FIRST_PDATE:2022) |
